# The role of comorbid childhood mental health conditions in the persistence of ADHD symptoms: Systematic review and Meta-analysis

**DOI:** 10.1101/2025.03.15.25324018

**Authors:** Yuan You, Tom McAdams, Yasmin I Ahmadzadeh, Tabea Schoeler, Filip Marzecki, Helena M.S Zavos

## Abstract

**Background:** Children diagnosed with ADHD and other comorbid mental health conditions often exhibit more severe functional impairments than those without comorbid conditions, including a tendency for their ADHD symptoms to persist into later developmental stages. We conducted a systematic review and quantitative analysis to investigate the extent to which specific childhood comorbidities (internalizing, externalizing and neurodevelopmental conditions) predict the persistence of childhood ADHD into later developmental stages.

**Methods:** We extracted data from 26 studies meeting the criteria for inclusion and applied multilevel random effects models to obtain pooled estimates of Cohen’s *d* for selected predictors on ADHD persistence.

**Results:** Childhood comorbid internalizing and externalizing conditions (*d*=0.19 and *d*=0.31, respectively), but not neurodevelopmental disorders, were significantly associated with ADHD persistence. After adjusting for covariates (sex, age and other comorbidities), this association diminished for externalizing conditions (*d*_adj_=0.24) and was no longer significant for internalizing conditions (*d*_adj_=0.05). The association between comorbid externalizing behavior problems and ADHD persistence was found only in studies that used parent-reported data to measure childhood ADHD and externalizing conditions, but not in studies that included teacher-reported childhood symptoms.

**Conclusions:** Childhood comorbid externalizing and, to a lesser extent, internalizing conditions were associated with the persistence of ADHD, but this association may be partially due to confounders. Childhood comorbidity of neurodevelopmental disorders does not appear to increase the likelihood of ADHD persistence.

## 1. Introduction

As a highly heritable neurodevelopmental disorder (Freitag et al., 2010; Larsson et al., 2014; Stevenson., 1992), Attention Deficit Hyperactivity Disorder (ADHD) often persists from childhood into adulthood (e.g. Caye et al., 2016; Roy et al., 2016). For those with childhood ADHD, between 60-80% will maintain functional impairment and 30% still meet the full diagnostic criterion in early adulthood (Biederman et al., 2010). Persistent ADHD is typically associated with greater functional impairment, including being associated with lower educational attainment, difficulties in the workplace (Fredriksen et al., 2014), and more severe mental health conditions (Yoshimasu et al., 2018). Common quantitative approaches to understanding ADHD persistence include measuring the stability of diagnosis (e.g., Biederman et al., 2011; Law et al., 2014; Mick et al., 2011) and measuring the stability of symptoms (e.g., Loya, 2012; Brown et al., 2022). In studies that assess the stability of ADHD diagnosis, persistence is defined as meeting diagnostic criteria both at baseline and during follow-up, typically determined through clinical interviews. In studies that assess the stability of ADHD symptoms, trajectory-based approaches characterize ADHD persistence by identifying stable and elevated symptom trajectories over time.

A broad spectrum of psychopathologies frequently co-occur with ADHD, including internalizing, externalizing, and neurodevelopmental conditions (e.g. Cuffe et al., 2020; Reale et al., 2017; Spencer, 2006). Children diagnosed with ADHD who also have comorbid mental health conditions often exhibit more severe functional impairments, including a tendency for their ADHD symptoms to persist into later developmental stages (Biederman et al., 1996, 2012; Caye et al., 2016). Therefore, it is crucial to identify individuals whose ADHD symptoms are likely to persist into later stages. Understanding the role of childhood comorbidities in ADHD persistence could, in the future, help to identify those whose ADHD symptoms are more likely to persist into adulthood and help guide interventions.

In studies defining ADHD persistence based on clinical diagnostic criteria, internalizing conditions have been consistently associated with the persistence of ADHD diagnosis (e.g. Caye et al.,2016; Law, Sideridis, Prock, & Sheridan, 2014). Children with high levels of depression symptoms or anxiety symptoms in questionnaires (e.g., Mick et al., 2011; Murry et al., 2022) or diagnosed with depression or anxiety through clinical interviews (e.g., Biederman et al., 2012; Chang et al., 2011; Cheung et al., 2015; Kessler et al., 2005) before the age of 12 are more likely to meet the diagnostic criteria for ADHD over a period ranging from 1 to 7 years. However, the association between internalizing conditions and ADHD persistence is not consistent across all studies employing a diagnostic definition of persistence. For example, a study found that major depressive disorder and multiple anxiety disorders were not associated with ADHD persistence after controlling for demographic confounders in an 11-year follow-up study with children who were 11 years old at the initial assessment (Biederman et al., 2012).

In studies defining ADHD persistence using trajectory-based approaches, no significant predictive effects of internalizing comorbidities have been identified in relation to the persistence of ADHD in latent growth models over a 10-year period, when controlling for age at baseline in children initially under 12 years old (Loya, 2012). However, in studies utilizing latent class models without including covariates, fewer depressive symptoms were associated with the declining ADHD group (Riglin et al., 2016). Therefore, previous studies do not provide a clear picture of the association between internalizing conditions and ADHD persistence.

Externalizing conditions, such as oppositional defiant disorder and conduct disorder, have also been associated with persistence of ADHD based on clinical diagnostic criteria from mid-childhood to adolescence or adulthood (e.g., Biederman et al., 2011; Cherkasova et al., 2013; Law et al., 2014). However, the association between externalizing conditions and ADHD persistence are also inconsistent. For instance, Gao et al. (2015) adjusted for factors such as living conditions, baseline ADHD symptoms, and other comorbidities and found that oppositional defiant disorders and conduct disorders were not associated with the persistence of ADHD diagnosis. Similarly, Kessler et al. (2005) found that after controlling for ADHD severity and treatment, comorbidities such as oppositional defiant disorders and conduct disorders at age 15 were not associated with the persistence of ADHD diagnosis into adulthood (18-44 years old).

Significant results have been reported in studies investigating the predictive effect of externalizing conditions on ADHD trajectories. Childhood externalizing comorbidities, such as aggression and oppositional defiant disorders have emerged as predictors of changes in ADHD symptoms throughout the developmental course from 9 years old to 20 years old (Loya, 2012). These results are further supported by studies using latent class analysis, indicating that individuals with higher levels of oppositional defiant disorders (Brown et al., 2022; Musser et al., 2016) and aggression (Sasse et al., 2016) before the age of 12 are more likely to persist with their ADHD symptoms in the 2 to 9 years following initial assessment. Overall, there have been numerous studies examining the significant associations between externalizing conditions and the persistence of ADHD diagnosis or trajectory. It appears possible that some of the variation in results may be due to differences in the analyses and particularly whether analyses were adjusted to control for the severity of ADHD symptoms and other covariates.

Studies that have focused on the relationship between ADHD and other neurodevelopmental conditions have not found a significant association with the persistence of ADHD based on clinical diagnosis. When using diagnosis as a criterion to define the persistence of ADHD, learning disorders (Biederman et al., 1996; Chang et al., 2011; Rinsky, 2012; Willcutt et al., 2007) and tic disorders (Chang et al., 2011) have not been linked to the persistence of ADHD. The only significant associations relate to intellectual disability. Children with intellectual disability have been identified as more likely to be in the early onset/stable ADHD group (Neece et al., 2011).

Studies which have focused on the developmental trajectories of ADHD, neurodevelopmental conditions such as autism (Stringer et al., 2020), and language difficulty (Yew & O’Kearney., 2017) have not been associations with the slope of ADHD trajectories when multivariate regressions or multivariate growth model were used. In these two studies, other variables that might influence the trajectory slope, such as other comorbidities and basic demographic variables (for example, SES, parenting quality, child temperament) were controlled for. However, similar to the results obtained using diagnostic criteria, intellectual disability has been shown to be associated with ADHD persistence in latent class analysis in a 13-year follow-up study that began when children were under 12 years old without controlling for other covariates (Riglin et al., 2016). Therefore, it appears that the specific type of neurodevelopmental disorder and the inclusion of covariates may influence study outcomes.

Furthermore, ADHD comorbidities are themselves influenced by covariates such as sex. For example, among females with ADHD, the predominantly inattentive presentation is more common (Gaub & Carlson, 1997; Nussbaum, 2012), and they tend to exhibit fewer externalizing problems (Mayes et al., 2020). However, females are generally more susceptible to internalizing problems such as anxiety and depression (Shorey et al., 2022; Jalnapurkar et al., 2018). Therefore, sex may serve as a covariate in the relationship between childhood comorbidities and ADHD persistence. Additionally, ADHD is a neurodevelopmental disorder that is closely linked to age and developmental stages (Holland & Sayal, 2019). As a result, the age of onset and period of follow-up may also be important covariates. Moreover, reporter bias may influence findings. For instance, studies have shown that parents are more likely to report persistent ADHD symptoms over time compared to individuals who self-report their ADHD symptoms (Barkley et al., 2016).

Recognizing the importance of understanding how comorbidities influence the persistence of ADHD symptoms, one previous meta-analysis has been published on this topic (Caye et al., 2016). In this meta-analysis, persistent ADHD was defined as meeting diagnostic criteria both before 12 years old and after 18 years old The study found that major depression, and oppositional defiant disorders were significantly associated with the persistence of ADHD symptoms. However, several gaps in knowledge still exist. For example, it remains unclear from this meta-analyses whether the associations between persistent ADHD and these co-occurring symptoms may be attributable to other socio-demographic variables that increase the risk for ADHD, emotional disorders, and externalizing disorders. In addition, more studies are have now been published in this area which will enable broader exploration of possible co-morbidities. For example, anxiety was not included in the meta-analysis conducted by Caye due to a lack of sufficient studies.

In this paper, we conducted a systematic review and meta-analysis to investigate whether specific childhood comorbidities (internalizing, externalizing and neurodevelopmental disorders) were associated with the persistence of childhood ADHD at any later developmental stage. Within the meta-analysis, we also investigated how covariates such as sex and baseline ADHD might moderate the results.

## 2. Method

### 2.1 Search Strategy

Our search strategy was registered using PROSPERO (protocol number: CRD42023442127). Studies included in this review were peer-reviewed articles written in English or Chinese. Two databases were searched: (1) Web of Science. Indexes = SCI-EXPANDED, SSCI, A&HCI, CPCI-S, CPCI-SSH, ESCI (Timespan=All years); (2) Ovid platform. Indexes = Embase (1974 - 2023), Ovid MEDLINE(R) (1946 - 2023), Global Health (1973 - 2019), APA PsycArticles Full Text, APA PsycINFO (1806 - 2023). The search terms primarily define our population of interest (i.e., the initial assessment was in childhood), the exposures of interest (e.g. psychopathologies), the type of study (longitudinal), and the outcome of interest (ADHD persistence). Detailed search terms are provided in the supplementary materials (see **Appendix S1**. Search strategies).

All searches were conducted independently by two of the authors, YY and FM, between September – December 2023. Any disagreements were settled by HZ and TM. Manual searches of published systematic reviews and the reference lists of included studies were also performed. Full screening was conducted on articles where the title and abstract met the inclusion criteria (see below). If we were unable to conclude whether the article met inclusion criteria during title and abstract screening, it underwent full-text screening process.

### 2.2 Study Selection

Longitudinal prospective studies were included if they collected data for both ADHD and comorbid psychopathologies in childhood (before age 12), as well as ADHD symptoms at later ages. Studies were included regardless of the time period over which data was collected. In order to investigate factors that could predict the persistence of ADHD symptoms, only studies where we were able to distinguish between persistent and remitted ADHD symptoms were included. Most studies adopted one of two ways to explore the persistence and remission of ADHD: (1) using a cut-off to distinguish persistent and remitted ADHD; (2) using trajectories to explore the development of ADHD symptoms. If studies used a cut-off to distinguish persistent and remitted ADHD, the persistence of ADHD was defined as meeting the threshold both in childhood and at the second time point. The diagnostic threshold was defined as either the criterion in questionnaires/clinical interviews or diagnostic assessment by a clinician. Where studies used trajectories to explore the development of ADHD symptoms, the persistence of ADHD was defined as individuals within the trajectory that showed consistently high levels of ADHD symptoms at each time point measured.

The PRISMA flowchart is presented in Figure 1. We initially screened 2000 studies from the database, and an additional 3 studies were added manually. After reviewing the abstracts and full texts, a total of 26 studies were included in our meta-analysis. The excluded studies primarily lacked sufficient measurement of ADHD and comorbidities during childhood or emotional problems at later stages. Other exclusion criteria included: meta-analyses, reviews, or case studies; studies primarily focused on ADHD treatment; non-peer-reviewed articles; animal studies; studies missing any of the following measurements: childhood or adulthood ADHD, or childhood comorbidities; studies focusing on children with specific comorbid physical conditions; studies focusing on special periods, such as the COVID-19 pandemic; studies that could not distinguish between persistent and remitted ADHD; and studies written in languages other than English and Chinese.

**Figure 1.**
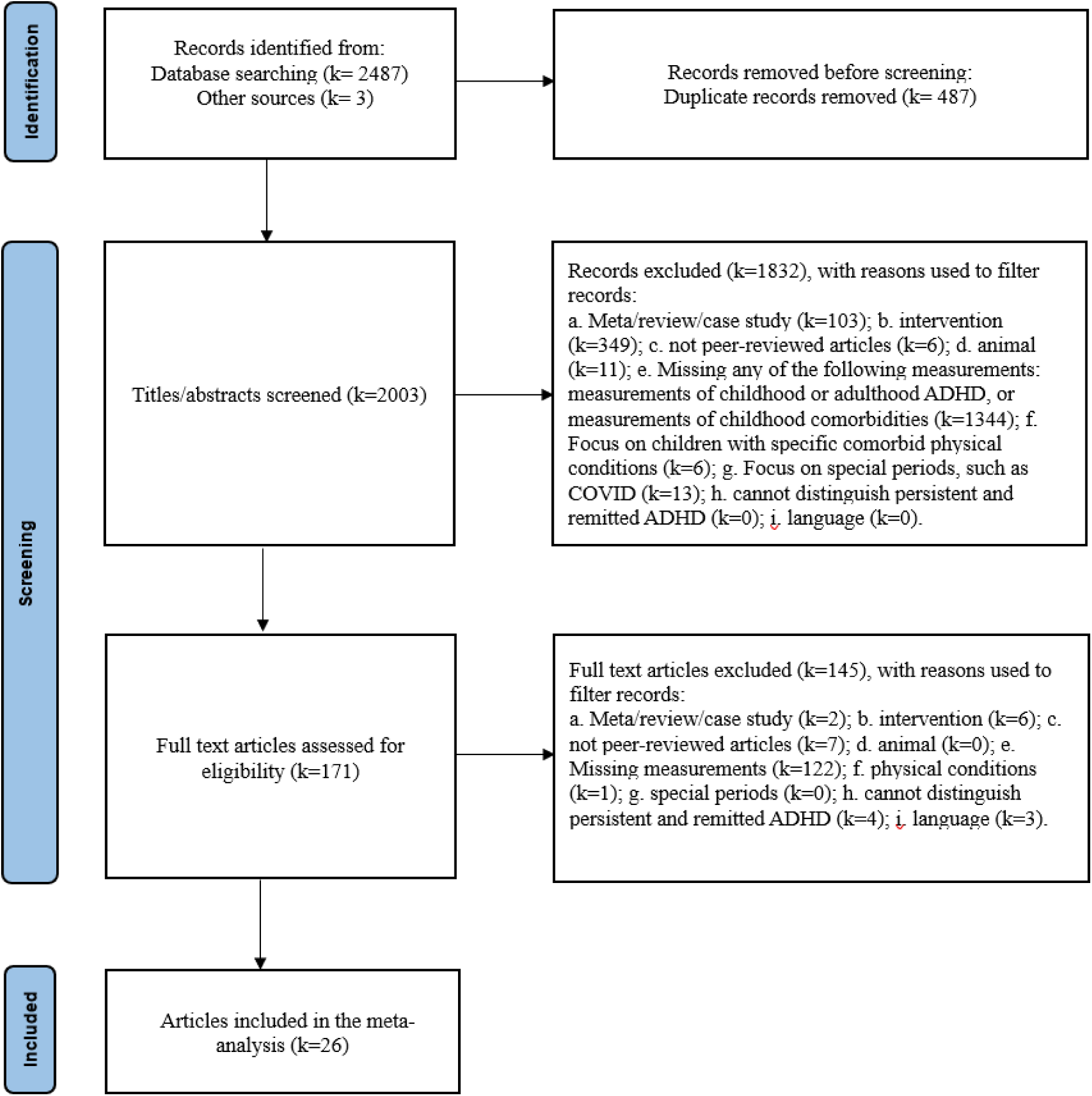
Flowchart.

Four studies used slightly different definitions of ADHD persistence than we had described in our pre-registration (Biederman et al., 1996; Biederman et al., 2011; Biederman et al., 2012; McAuley et al., 2017). We included them in our initial analysis and conducted a sensitivity analysis excluding them to determine their impact on our findings. Among them, three studies included broader ADHD persistence than we initially proposed in our pre-registration that defined persistence as either fulfilling diagnostic criteria, meeting sub-threshold diagnostic criteria (exhibiting more than half of the full diagnostic symptoms), experiencing functional impairment, or having taken ADHD medication. As a result, these studies may include more participants in the ADHD persistence group. In another study (McAuley et al., 2017), the predictive effect of adjusted ODD on ADHD persistence was determined by comparing a fully persistent group (meeting more than six DSM-IV criteria for ADHD symptoms and having a Children’s Global Assessment Scale score over 60) with a fully remittent group only (meeting fewer than six DSM-IV criteria for ADHD symptoms and having a Children’s Global Assessment Scale score below 60). In this study, the definition of ADHD persistence and remission is stricter than we planned in our pre-registration. Because these four studies provide information relevant to our research question, we included them in our initial analysis and conducted a sensitivity analysis excluding them to determine their impact on our findings.

### 2.3 Data Extraction

Most studies collected data using clinical assessment tools, so comorbidities were treated as binary variables in these studies. Results were typically reported as odds ratios (OR), which indicate the relative likelihood that individuals with persistent ADHD had childhood comorbidities compared to those with remitted ADHD.

For studies, which measured childhood comorbidities using quantitative questionnaires, continuous scores of comorbidities in both persistent and remitted ADHD groups were reported. For these studies we extracted the mean and standard deviation of childhood comorbidities in persistent ADHD and remitted ADHD groups and converted these to Cohen’s *d*. In situations where *p*-values were not reported as numerical data, we chose the most conservative way to derive Cohen’s *d*. More specifically, we set *p*=1 (i.e., Cohen’s *d* = 0) whenever non-significant results were reported as *p*>0.05, and we set *p*=0.05 whenever significant results were reported as *p*<0.05.

### 2.4 Multilevel Random Effects Model

The R package *metaphor* (Viechtbauer, 2010) was used to conduct the multilevel random effects models (MREM). MREMs were used to allow for the inclusion of dependent effect sizes, e.g., when pooling together multiple estimates that were obtained from the same underlying sample (such as the effect sizes for comorbidities reported by different reporters, and effect sizes before and after adjusting for covariates). We considered four sources of variation in the MREMs (see **Figure 2**): Level 1, the sampling variance of all extracted effect sizes; Level 2, the variance within the same cohort and same comorbidity type (measurement differences); Level 3, the variance in effect sizes between different comorbidity types within the same cohort; and Level 4, the variance between different cohorts. Log-likelihood ratio tests (Assink & Wibbelink, 2016) were used to test the significance of heterogeneity in Levels 2, 3, and 4. The *I*^2^ statistic was used to test the heterogeneity between effect sizes (Higgins et al., 2003). *I*^2^ indicates what proportion of variation among studies included in the meta-analysis is due to heterogeneity. An *I*² less than 25% indicates low heterogeneity; an *I*² between 25% and 75% indicates moderate heterogeneity; and an *I*² above 75% indicates high heterogeneity.

**Figure 2.**
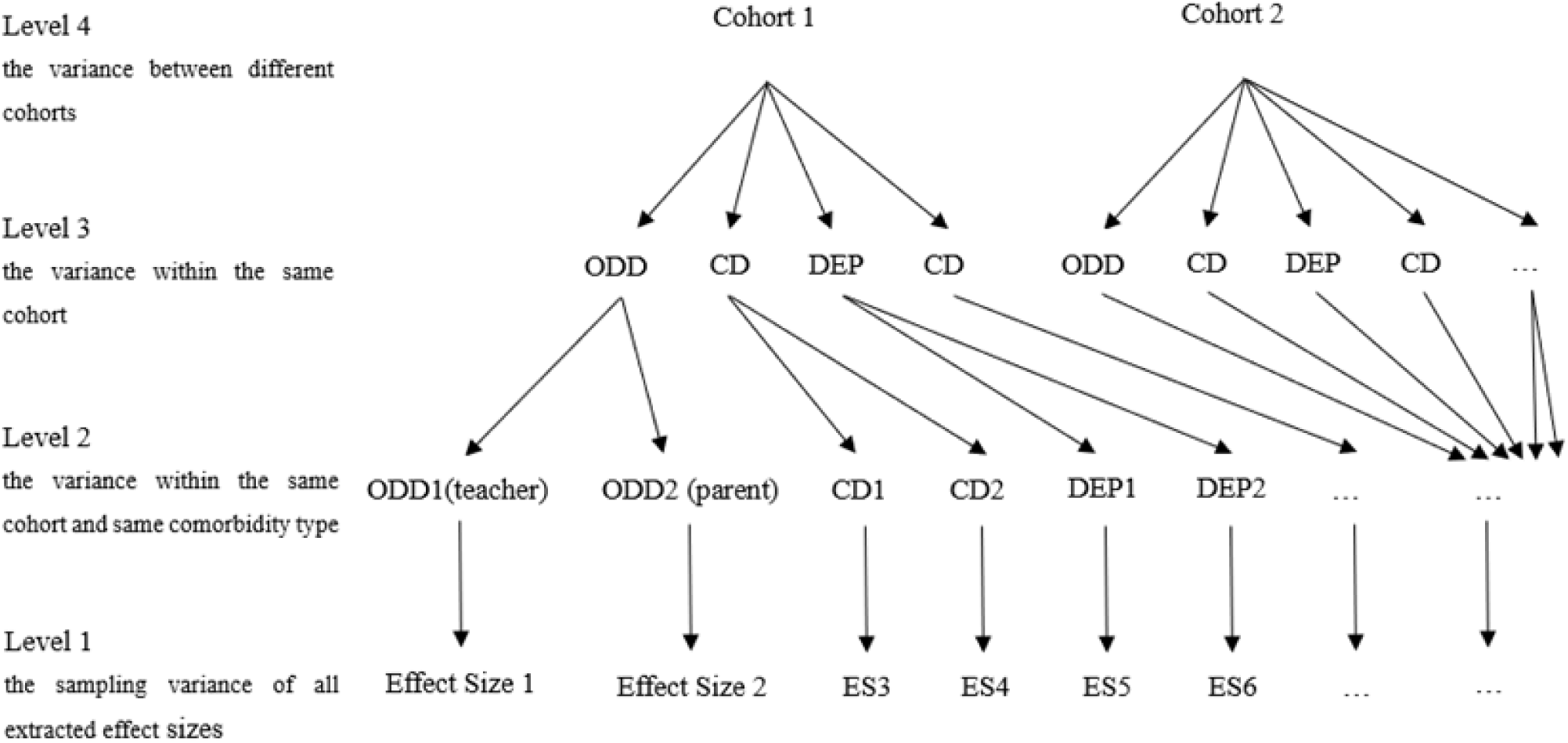
Four-level meta-analytic data structure of our study.

### 2.5 Moderators

We explored the moderating effects of sex, age, measurement and reporter on estimates in the current study. The Omnibus test of moderators (QM) was used to test the potential moderators in the multilevel random effects model. If the p-value for QM is significant, it indicates that the moderating variable explains the heterogeneity in the model (Assink & Wibbelink, 2016).

### 2.6 Quality of studies and publication bias

Quality assessment for each study was conducted by YY using the NIH Quality Assessment Tool for Observational Cohort and Cross-Sectional Studies (NIH, 2014). In this tool, the research questions, selection of the study population, research design, and the validity and feasibility of the measurements are assessed. Based on the proportion of “yes” responses to the items in this tool, studies were categorized into four tiers: “good,” “fair,” “poor,” and “very poor.” Studies with 75%-100% “yes” responses were rated as good, typically indicating a low risk of publication bias. Studies with 50%-74% “yes” responses were rated as fair. Those with less than 50% “yes” responses were rated as poor (25%-49%) or very poor (0%-25%), indicating a higher risk of publication bias.

We further used multilevel Egger’s tests (Egger et al., 1997) and funnel plots (Sterne et al., 2001) to detect publication bias. Support for the null hypothesis in Egger’s test or symmetry in the plot would indicate the absence of publication bias.

## 3. Results

### 3.1 Study description

A total of 26 studies met the inclusion criteria, with publication dates ranging from 1983 to 2022, encompassing 24 distinct cohorts (see **Figure 1**, **Table 1, and Table 2**). These studies yielded 73 unadjusted outcomes and 47 adjusted outcomes. Specifically for internalizing conditions, we collected 14 adjusted results from 8 cohorts and 26 unadjusted results from 12 cohorts. Regarding externalizing conditions, our data comprised 30 adjusted results from 15 cohorts and 35 unadjusted results from 18 cohorts. For neurodevelopmental conditions, we obtained 2 adjusted results from 2 studies and 12 unadjusted results from 7 cohorts.

**Table 1.**
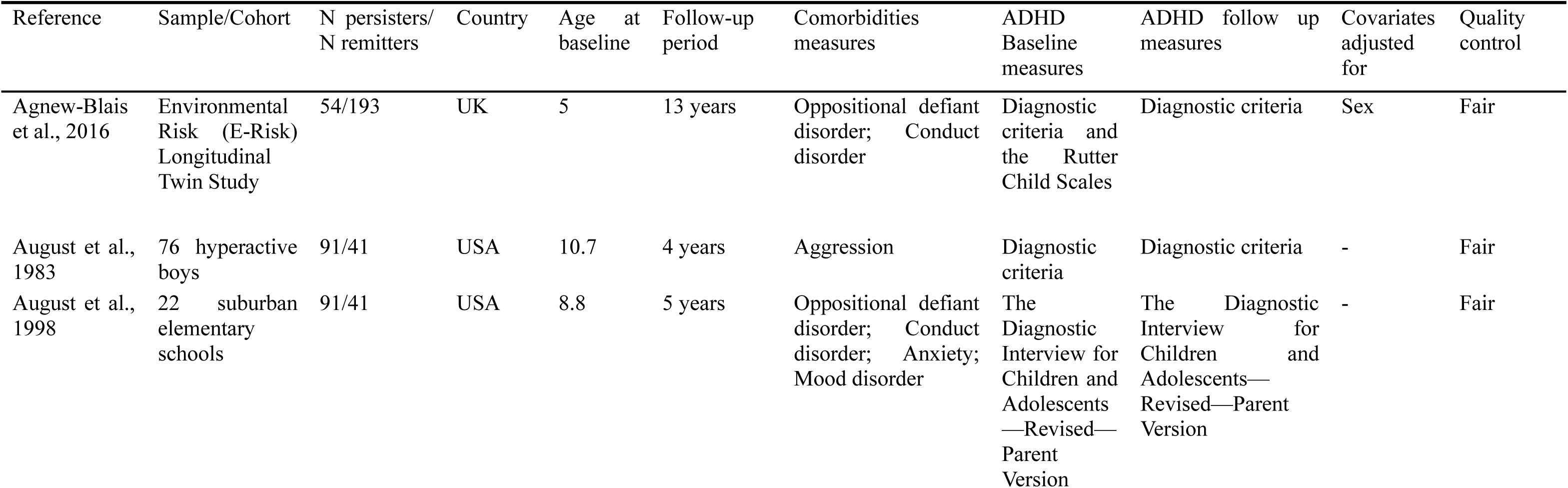
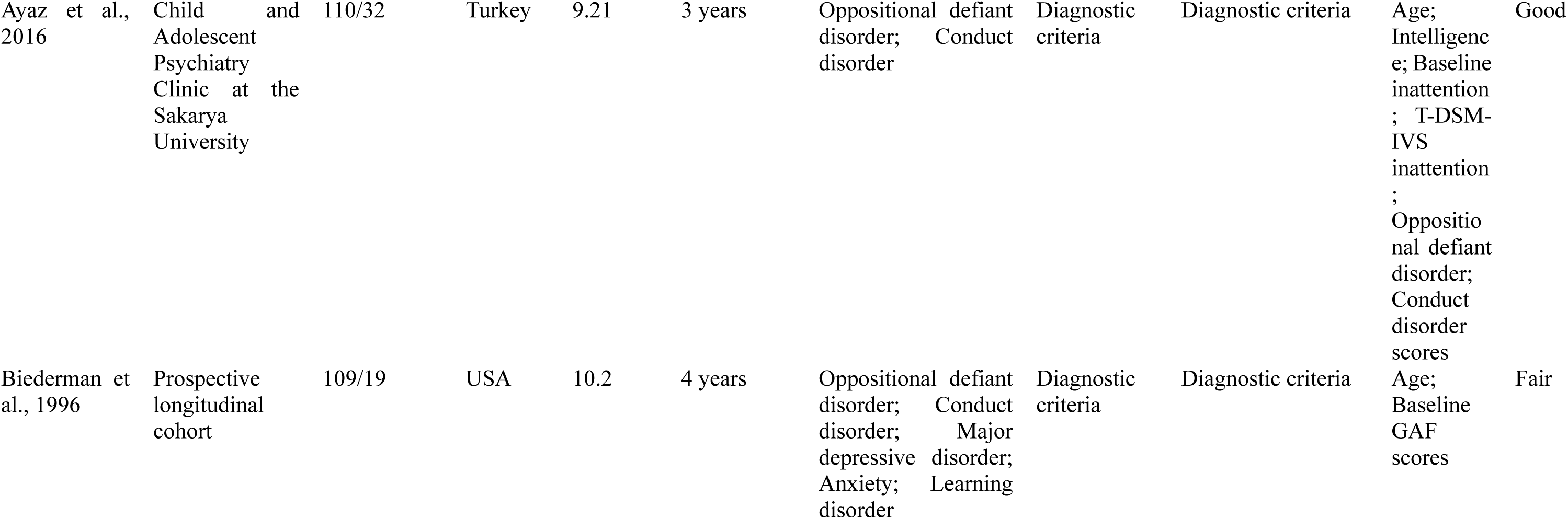
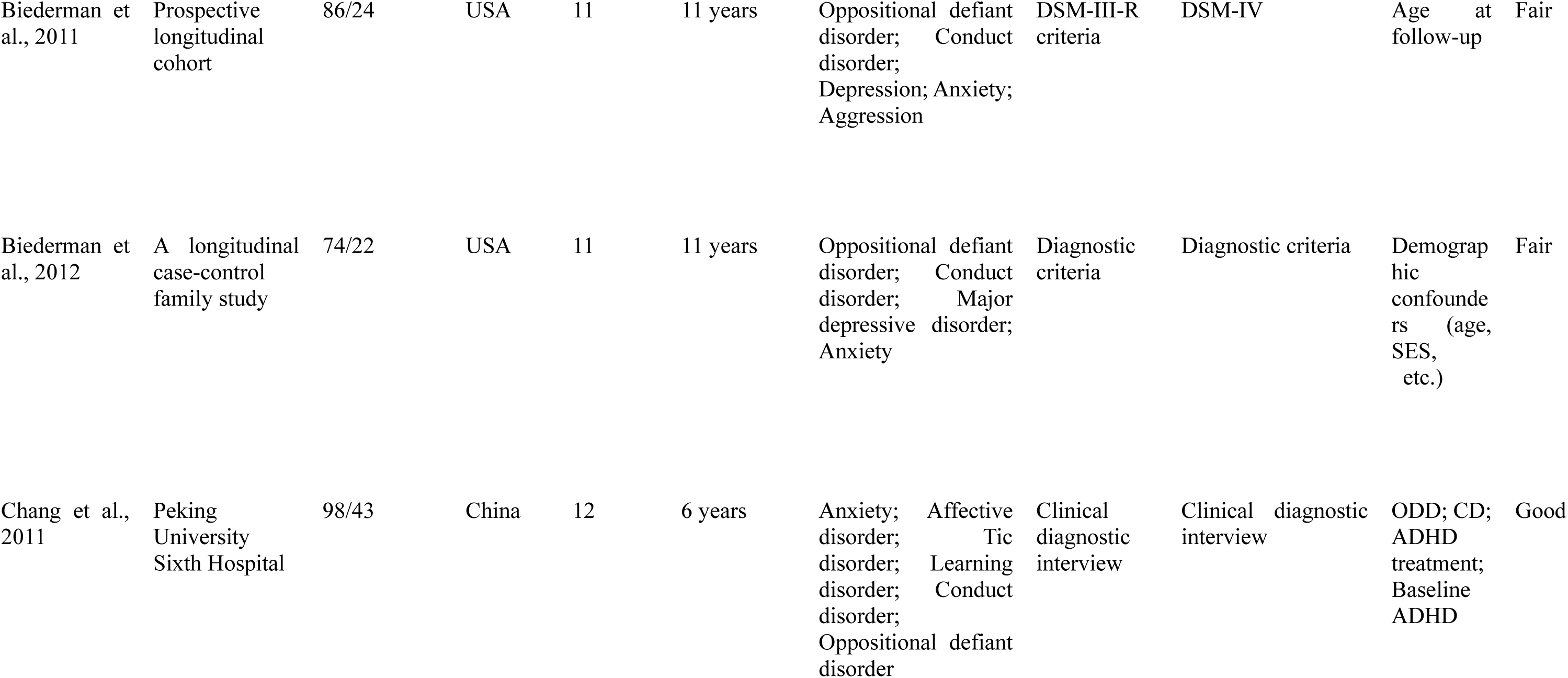
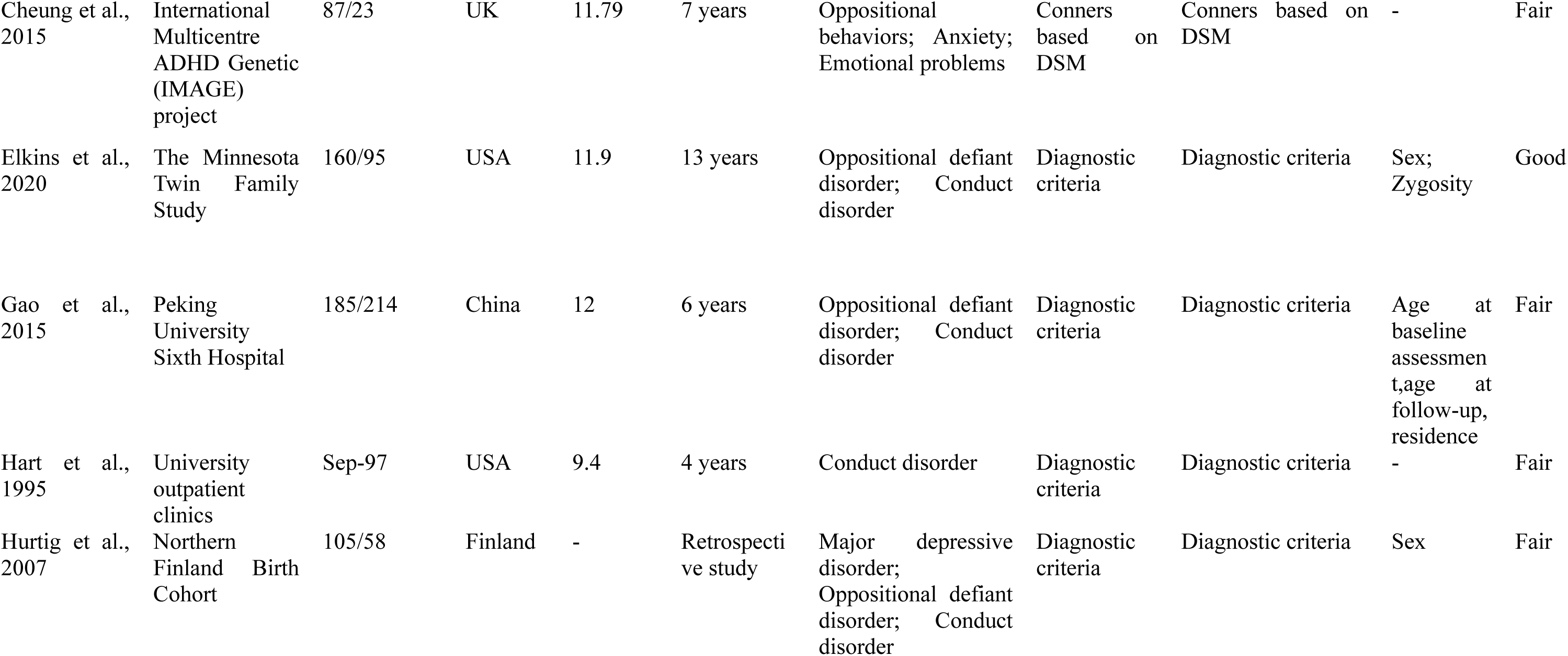
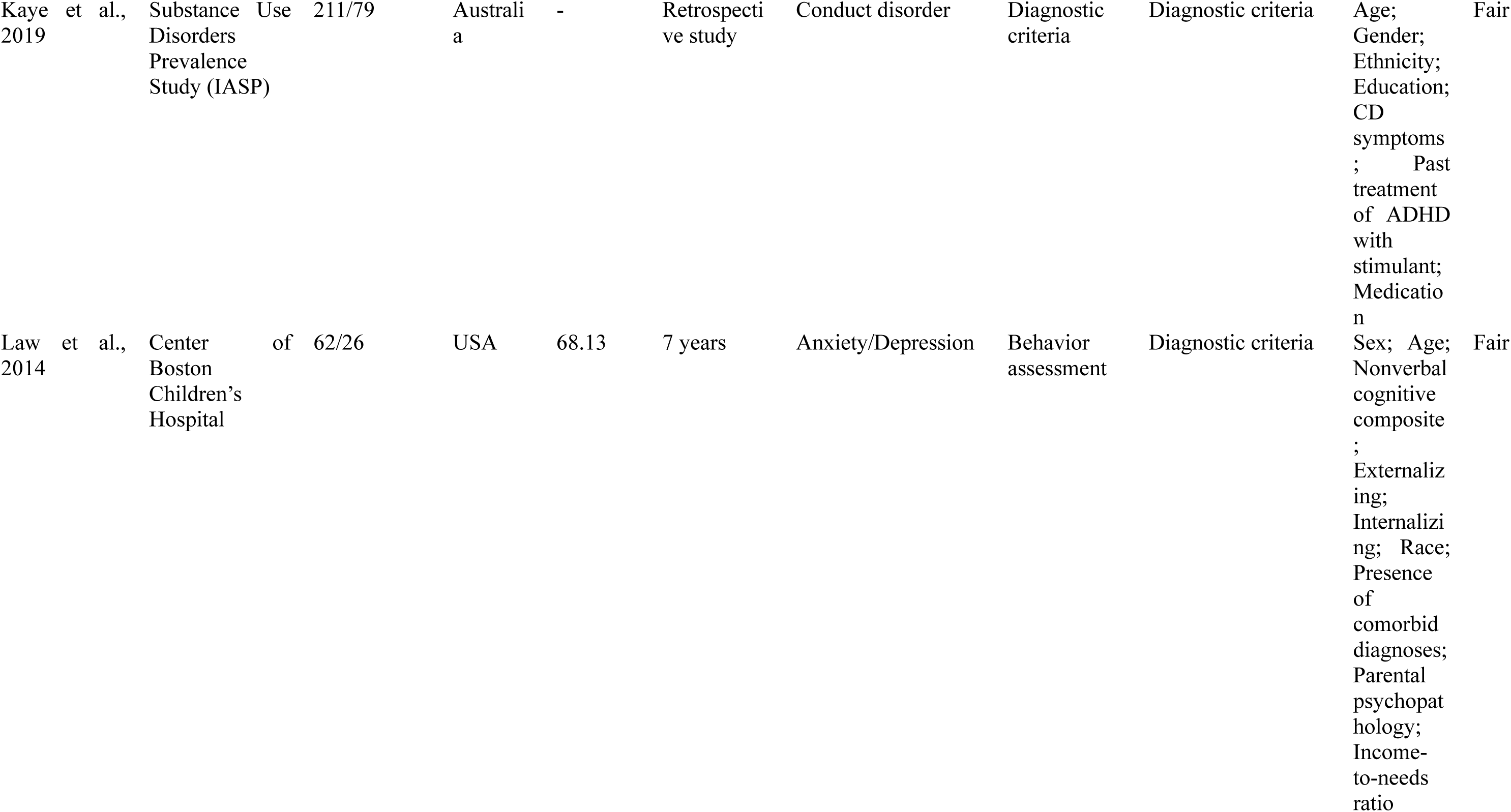
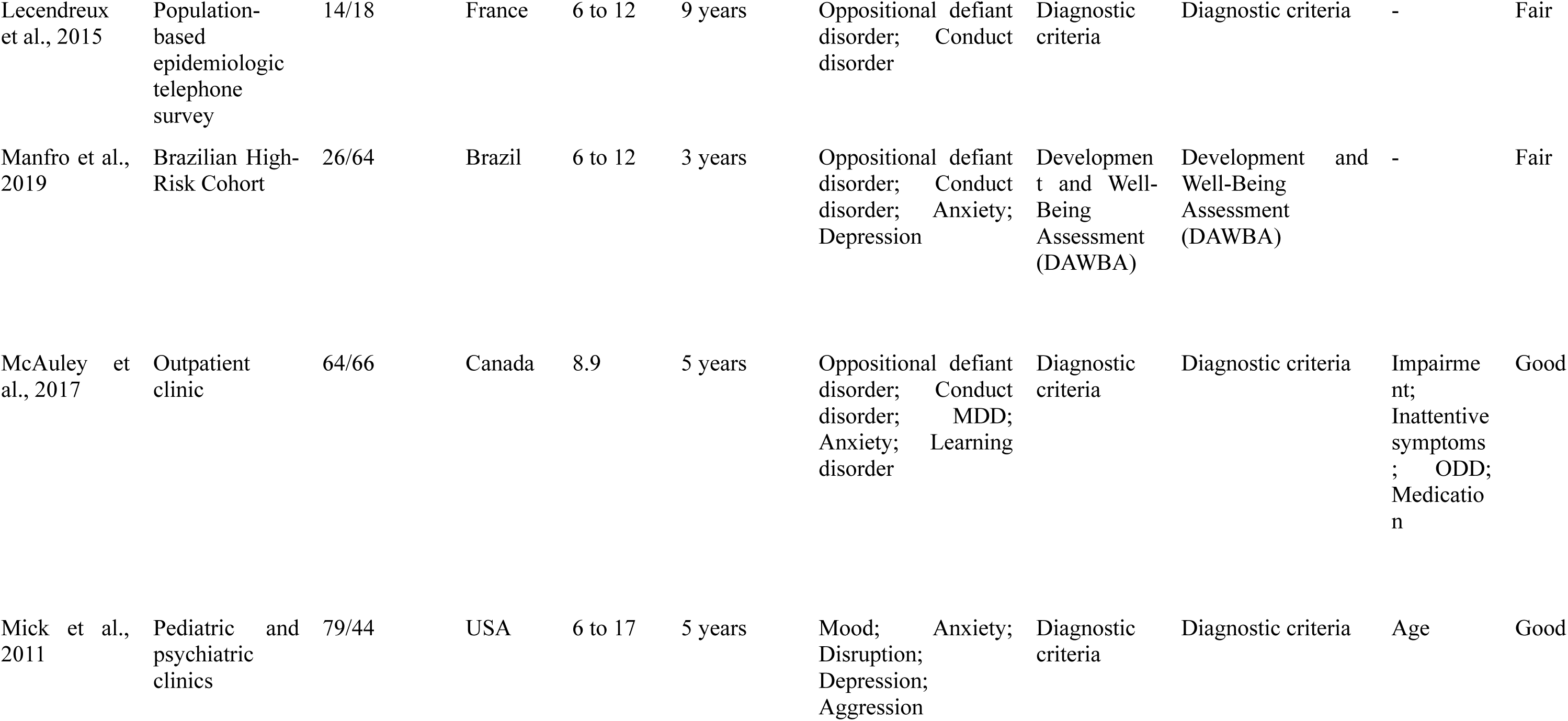
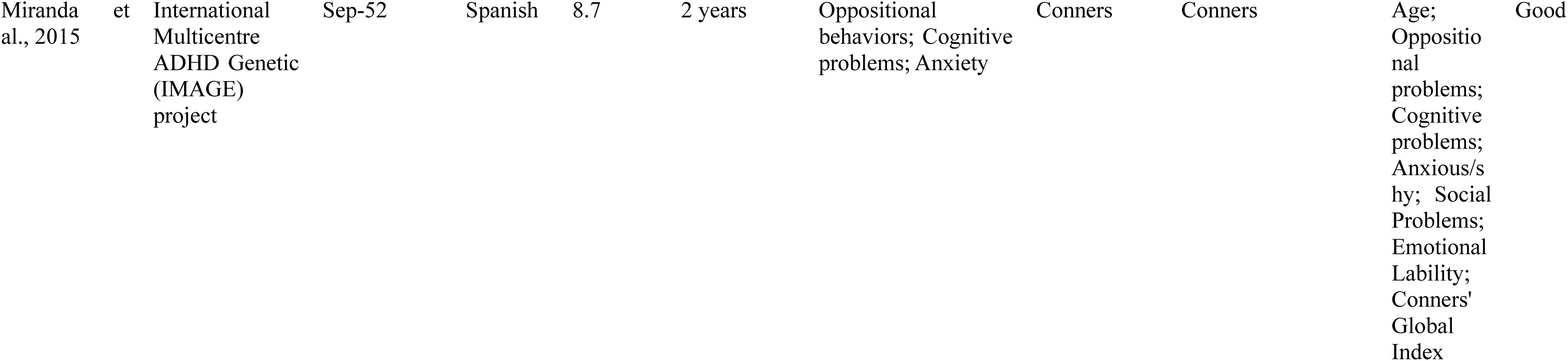
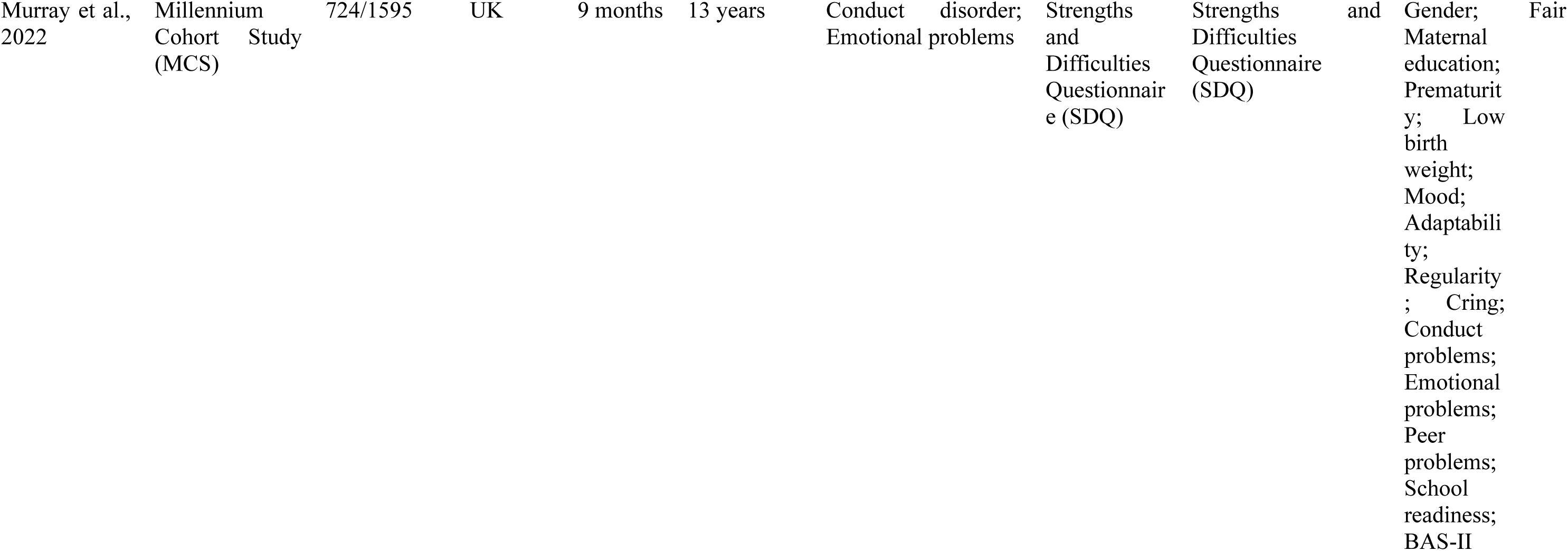
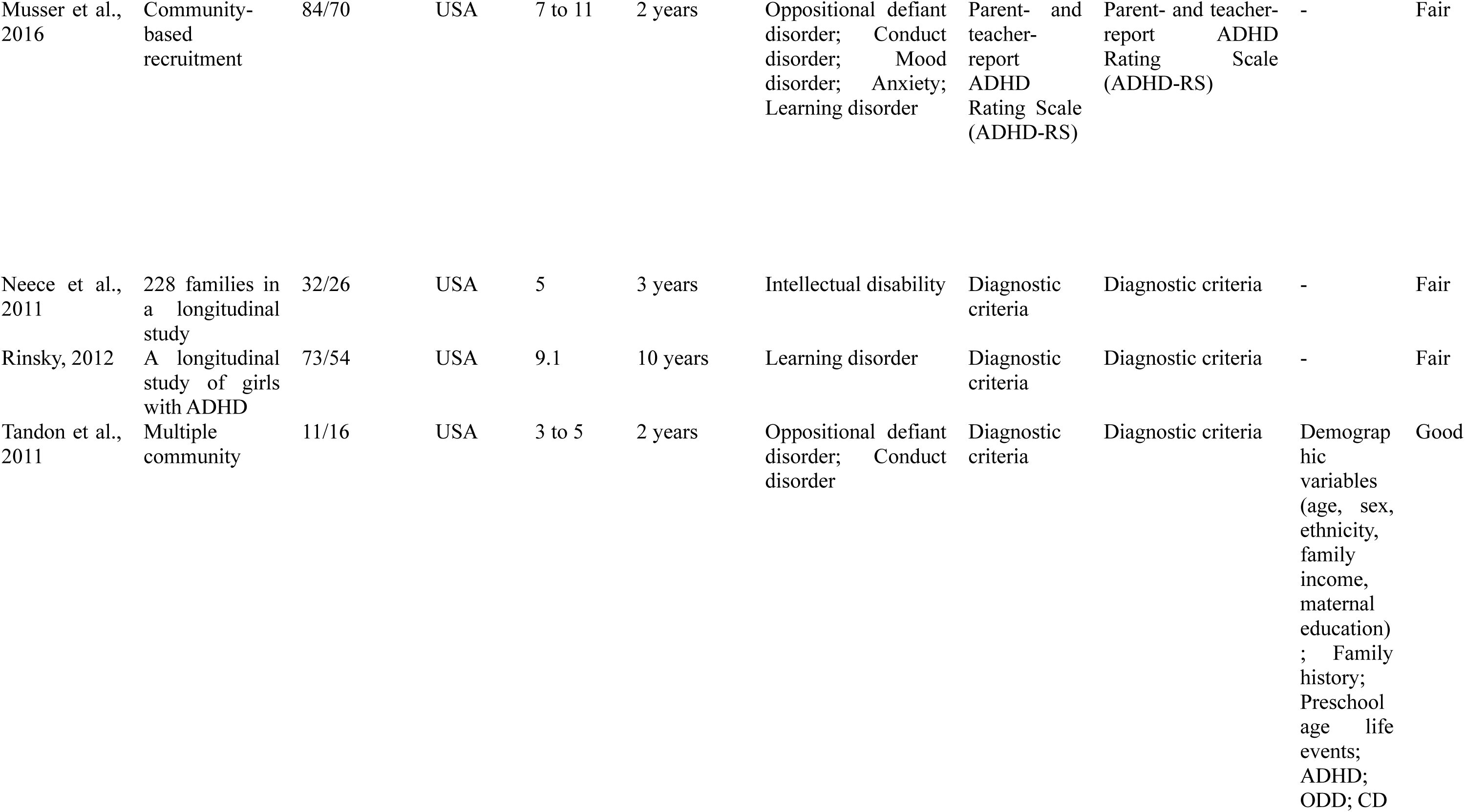
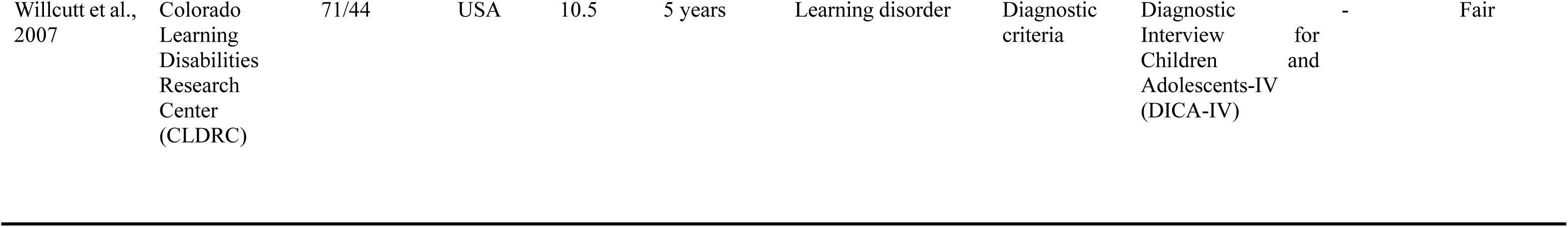
Studies investigating the predictive effect of comorbidities on ADHD persistence.

**Table 2.** Three-Level Random Effects Models: Associations between internalizing, externalizing, neurodevelopmental conditions and ADHD persistence.

| Models | Internalizing conditions | Externalizing conditions | Neurodevelopmental conditions |
| --- | --- | --- | --- |
| LRT<sub>Level 2</sub> | $\chi^2=0.00, p=1.00$ | $\chi^2=8.27, p=0.004$ | $\chi^2=0.00, p=1.00$ |
| LRT<sub>Level 3</sub> | $\chi^2=1.12, p=0.29$ | $\chi^2=0.00, p=1.00$ | $\chi^2=1.57, p=0.21$ |
| LRT<sub>Level 4</sub> | $\chi^2=0.00, p=1.00$ | $\chi^2=1.41, p=0.24$ | $\chi^2=0.00, p=1.00$ |
| $I^2$ | 33.88% | 62.26% | 48.30% |
| $I^2_{\text{Level 1\%}}$ | 69.13% | 16.26% | 46.58% |
| $I^2_{\text{Level 2\%}}$ | <0.01% | 50.76% | <0.01% |
| $I^2_{\text{Level 3\%}}$ | 30.87% | <0.01% | 53.42% |
| $I^2_{\text{Level 4\%}}$ | <0.01% | 32.98% | <0.01% |
| Unadjusted MREM models |  |  |  |
| $k_{\text{cohort}}$ | 12 | 18 | 7 |
| $k_{\text{cohort/comorbidities-type}}$ | 21 | 27 | 11 |
| $k_{\text{effect sizes}}$ | 26 | 35 | 12 |
| $d_{\text{pooled}}(95\% \text{ CI})$ | 0.19(0.08,0.30) | 0.31(0.19,0.43) | 0.13(-0.09,0.34) |
| $I^2$ | 15.36% | 56.00% | 50.25% |
| Adjusted MREM models |  |  |  |
| $k_{\text{cohort}}$ | 8 | 15 | 2 |
| $k_{\text{cohort/comorbidities-type}}$ | 14 | 26 | 2 |
| $k_{\text{effect sizes}}$ | 14 | 30 | 2 |
| $d_{\text{pooled}}(95\% \text{ CI})$ | 0.06(-0.06,0.18) | 0.24(0.11,0.37) | - |
| $I^2$ | 28.56% | 52.36% | - |

**Note:** $I^2_{\text{Level 1\%}}$ , proportion of variance due to the sampling variance of all extracted effect sizes; $I^2_{\text{Level 2\%}}$ , proportion of variance within the same cohort and comorbidity type; $I^2_{\text{Level 3\%}}$ , proportion of variance in effect sizes between different comorbidity types within the same cohort; and Level 4, proportion of variance between different cohorts. LRT, statistics from likelihood-ratio test.

The majority of studies were prospective and longitudinal. Initial measurements were typically taken between the ages of 5 and 12, with follow-up durations spanning 2 to 13 years. There was one retrospective study (Hurtig et al., 2007; Kaye et al., 2019), conducted with an adult sample.

### 3.2 Multilevel Random Effects Model: Effects of childhood comorbidities

#### 3.2.1 Internalizing conditions

Moderate heterogeneity (33.88%) across studies was found for the association between internalizing conditions and ADHD persistence (see **Table 2**). 69.13% of the variance was due to the sampling variance of all extracted effect sizes, while 30.87% was attributed to the differences in effect sizes between various comorbidity types within the same cohort. This suggests that different types of internalizing conditions may be associated with ADHD persistence in different ways.

As indicated in **Figure 3a** and **Figure 3b**, regarding internalizing conditions, only the unadjusted results instead of adjusted results suggested an association between internalizing conditions and ADHD persistence, with pooled Cohen’s *d* of 0.19 (0.08, 0.30) and 0.06 (−0.06, 0.18), respectively. Specifically, anxiety was significantly associated with the persistence of ADHD when no confounders were adjusted for, with *d*=0.20 (0.07, 0.34), but depression was not associated with ADHD persistence, with *d*=0.20 (−0.04, 0.44). Upon controlling for confounders, such as maternal education, gender (Murry et al., 2022), age (Miranda et al., 2015; Biederman et al., 1996), and baseline global assessment of functioning scores (Biederman et al., 1996), neither depression nor anxiety was associated with ADHD persistence (see Figure S1a, Figure S1b, Figure S1c, Figure S1d).

**Figure 3a.**
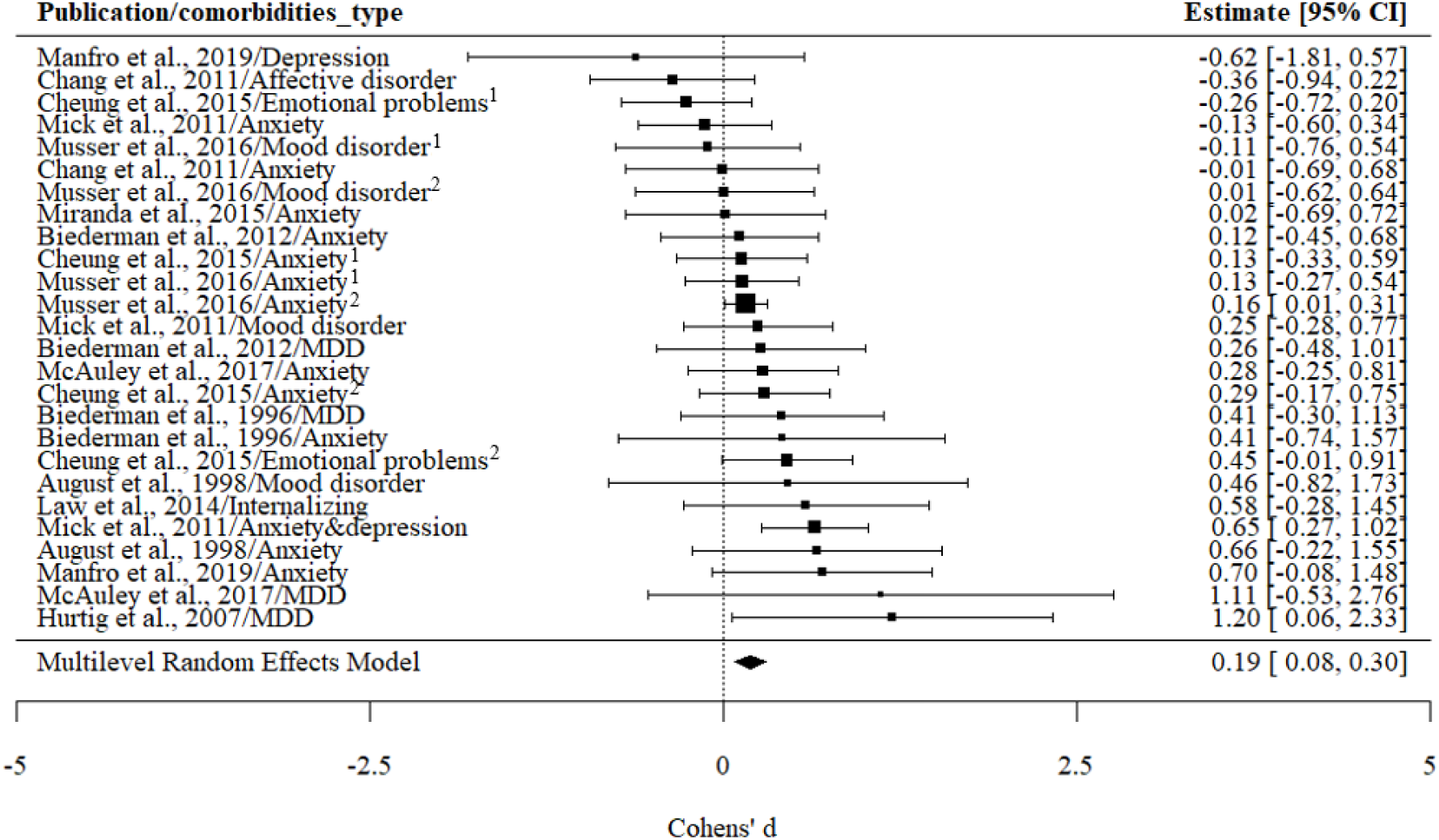
The association between internalizing and ADHD persistence: Unadjusted results. **Note**: Cheung et al., 2015/Emotional problems^1^: emotional problems were reported by teachers; Cheung et al., 2015/Emotional problems^2^: emotional problems were reported by parents. Cheung et al., 2015/Anxiety^1^: anxiety was reported by teachers; Cheung et al., 2015/Anxiety^2^: anxiety was reported by parents; Musser et al., 2016/Anxiety^1^: anxiety was reported by teachers; Musser et al., 2016/Anxiety^2^: anxiety was reported by parents. Musser et al., 2016/Mood disorder^1^: mood disorder was reported by teachers; Musser et al., 2016/Mood disorder^2^: mood disorder was reported by parents.

**Figure 3b.**
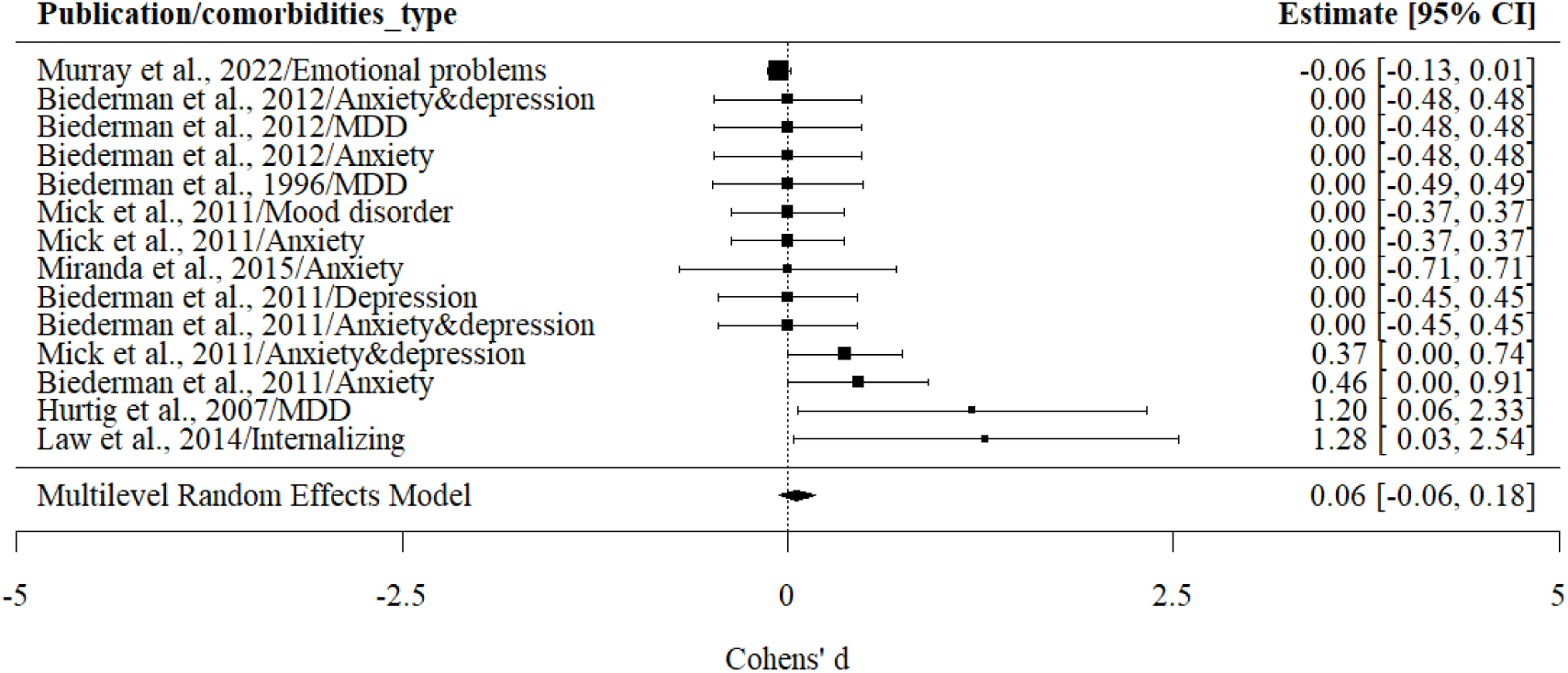
The association between internalizing and ADHD persistence: Adjusted results.

#### 3.2.2 Externalizing conditions

As is shown in Table 2, there was moderate heterogeneity among the association between externalizing conditions and ADHD persistence (*I*²=62.26%). Most of the variance between effect sizes was due to measurement differences within the same cohort and same comorbidity type (*I*²=50.76%). 32.98% of the variance was attributed to differences between cohorts, while the remaining 16.26% was attributed to the sampling variance of all extracted effect sizes. Therefore, differences in covariates within cohorts and differences in samples and measurement methods between cohorts may affect the association between externalizing conditions and ADHD persistence.

For externalizing conditions (see **Figure 4a** and **Figure 4b**), a larger pooled Cohen’s *d* (effect size) of 0.31 (0.19, 0.43) was observed for unadjusted results, compared to studies providing adjusted results, *d*_adj_=0.24 (0.11, 0.37). An attenuation in Cohen’s *d* was noted for both oppositional defiant disorder and conduct disorder when controlling for other covariates. The effect sizes were 0.28 (0.12, 0.44) and 0.25 (0.09, 0.41) for unadjusted results, respectively; 0.25 (0.07, 0.43) and 0.10 (0.06, 0.13) for adjusted results, respectively. Two studies reported unadjusted results for comorbid aggression on ADHD persistence (Mick et al., 2011; August et al., 1983). Only one of the two studies that did investigate the effect of aggression found an association with ADHD persistence (Mick et al., 2011). The pooled Cohen’s *d* of the adjusted results for the impact of comorbid aggression on the persistence of ADHD was 0.39 (−0.03, 0.80), indicating a non-significant effect either (see Figure S2a, Figure S2b, Figure S2c, Figure S2d, Figure S2e).

**Figure 4a.**
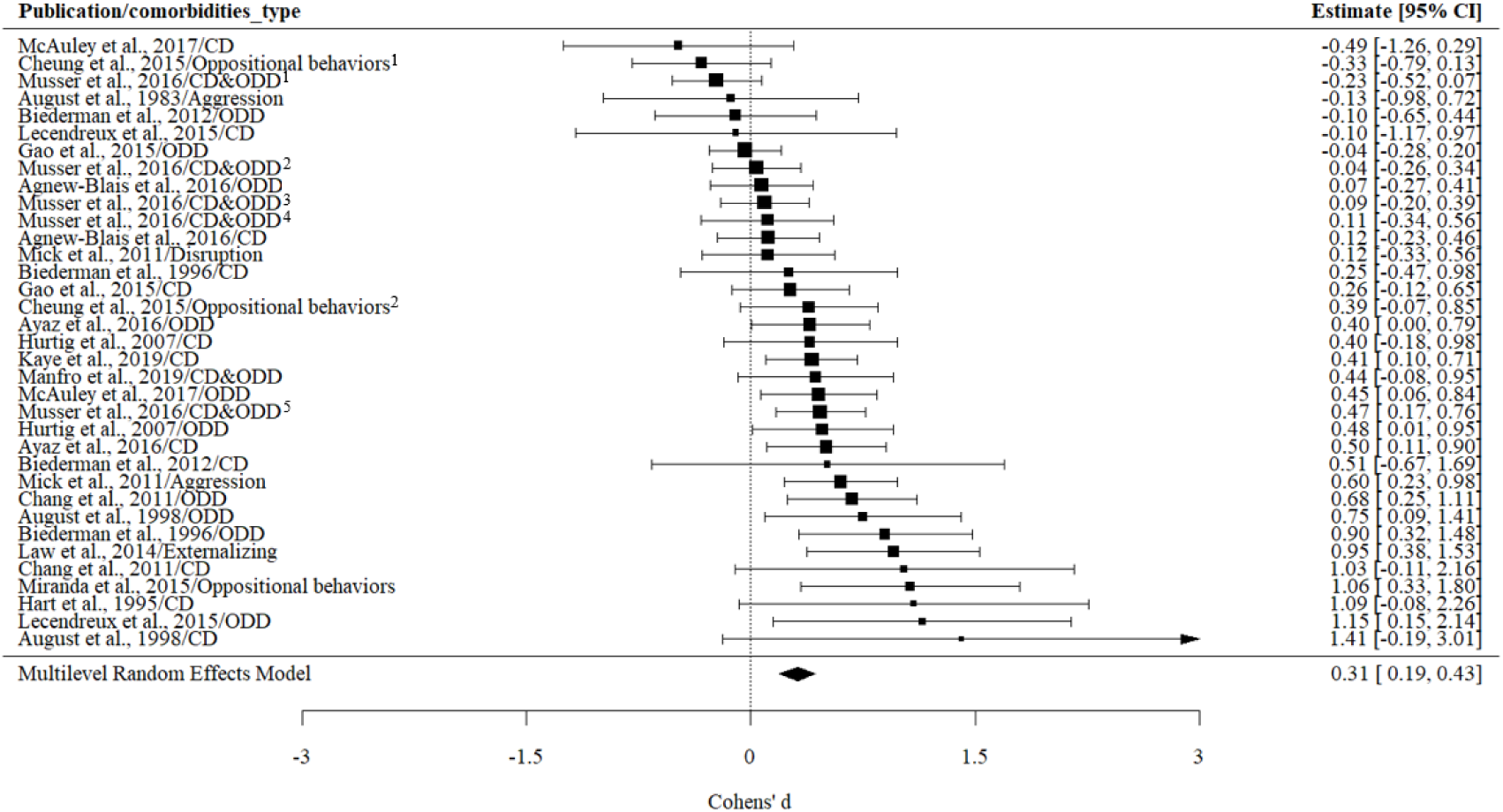
The association between externalizing conditions and ADHD persistence: Unadjusted results. Note: Cheung et al., 2015/Oppositional behaviors^1^: oppositional behaviors were reported by teachers; Cheung et al., 2015/Oppositional behaviors^2^: oppositional behaviors were reported by parents. Musser et al., 2012/CD&ODD^1^: CD&ODD was reported by teachers, and ADHD was reported by teachers. Musser et al., 2012/CD&ODD^2^: CD&ODD were reported by teachers, and ADHD was reported by parents. Musser et al., 2012/CD&ODD^3^: CD&ODD were reported by parents, and ADHD was reported by teachers. Musser et al., 2012/CD&ODD^4^: CD&ODD were assessed by diagnostic team, and ADHD was reported by teachers. Musser et al., 2012/CD&ODD^5^: CD&ODD were reported by parents, and ADHD was reported by parents.

**Figure 4b.**
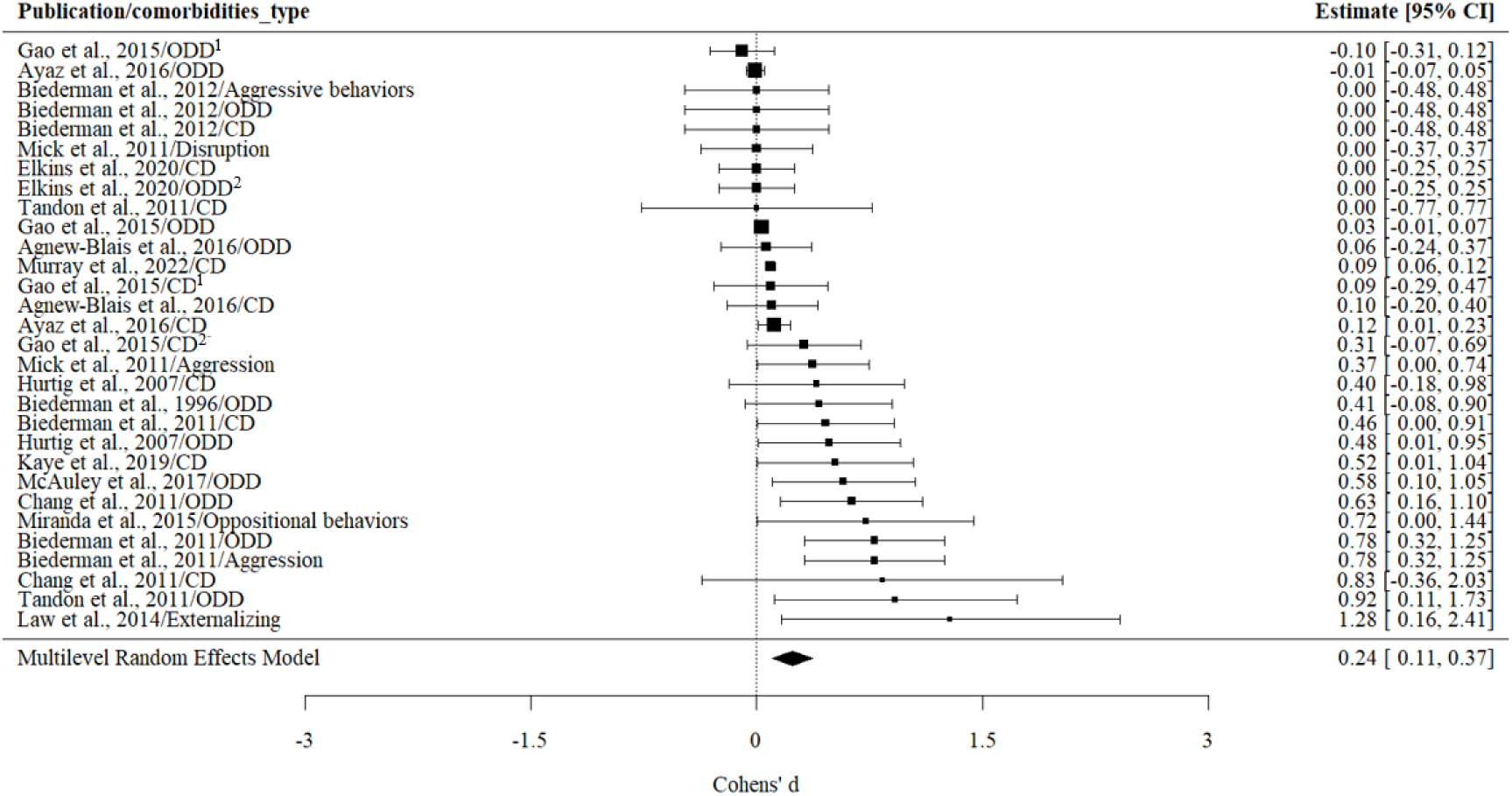
The association between externalizing and ADHD persistence_adjusted. **Note:** Gao et al., 2015/ODD^1^: ODD was included as a categorical indicator; Gao et al., 2015/ODD^2^: ODD was included as a quantitative trait. Gao et al., 2015/CD^1^: CD was included as a categorical indicator; Gao et al., 2015/ODD^2^: ODD was included as a quantitative trait.

#### 3.2.3 Neurodevelopmental conditions

There was moderate heterogeneity among effect sizes for the association between neurodevelopmental conditions and ADHD persistence (*I*^2^=48.30%) (see **Table 2**). Specifically, an important part of the heterogeneity was attributed to the sampling variance of all extracted effect sizes (*I*^2^=46.58%). The remaining variance was explained by the variance in effect sizes between different comorbidity types within the same cohort (*I*^2^=53.42%), suggesting difference in the association between specific neurodevelopmental conditions and ADHD persistence.

No significant association were found for overall neurodevelopmental conditions and ADHD persistence in unadjusted results (see **Figure 5a**), with a Cohen’s *d* of 0.13 (−0.09, 0.34). Specifically, childhood learning disorders was not associated with the persistence of ADHD, *d*=0.04 (−0.15, 0.23) (see Figure S3). The adjusted results regarding the association between neurodevelopmental conditions and ADHD persistence did not indicate significant findings (Biederman et al., 1996; Miranda et al., 2015).

**Figure 5a.**
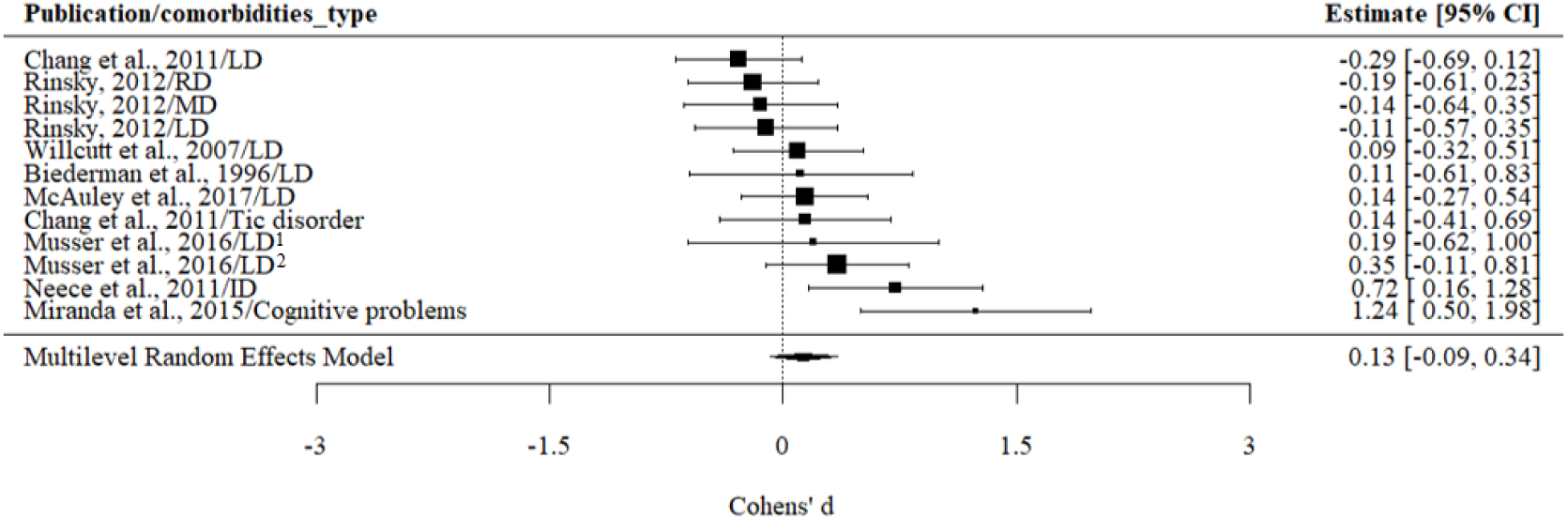
The association between neurodevelopmental conditions and ADHD persistence: Unadjusted results. **Note:** Musser et al., 2016/LD^1^: LD was reported by teachers; Musser et al., 2016/LD^2^: LD was reported by parents.

### 3.3 Multilevel Mixed Effects Model: Sources of Heterogeneity

To assess whether differences between unadjusted and adjusted results were significant, we used “whether the result is adjusted” as a moderator. It is found that the adjusting for covariates did not significantly moderate the association of either childhood internalizing (QM=2.39, *p*=0.12) or externalizing (QM=1.95, *p*=0.16) problems with ADHD persistence. (see Table 3).

**Table 3.** Source of heterogeneity.

| Moderators | Internalizing conditions |  | Externalizing conditions |  | Neurodevelopmental disorders |  |
| --- | --- | --- | --- | --- | --- | --- |
|  | QM | *p* | QM | *p* | QM | *p* |
| Whether the results were adjusted | 2.39 | 0.12 | 1.95 | 0.16 | 0.00 | 0.98 |
| Assessment method | 0.01 | 0.91 | 0.35 | 0.55 | 3.16 | 0.08 |
| Sex | 0.28 | 0.96 | 6.71 | 0.08 | - | - |
| Age at start | 1.01 | 0.31 | 1.96 | 0.16 | - | - |
| Tracking period | 4.18 | 0.12 | 4.71 | 0.09 | 10.60 | <0.05 |
| Reporters(childhood) | 0.06 | 0.97 | 24.00 | <0.05 | 6.39 | <0.05 |
| Reporters(adulthood) | 0.59 | 0.74 | 4.28 | 0.12 | - | - |

**Note:** There were not enough studies to test the moderating effect of sex and participant age at the start of study on the association between neurodevelopmental conditions and ADHD persistence.

The reporters of childhood ADHD and comorbid symptoms significantly moderated the association between childhood comorbid externalizing conditions and ADHD persistence (QM=24.00, *p*<0.05), but not the association between childhood comorbid internalizing conditions (QM=0.06, *p*=0.97) and ADHD persistence. Specifically, in the association between ADHD persistence and externalizing behavior problems, a significant result was observed only in studies using parent-reported childhood ADHD and comorbid externalizing conditions (*d*=0.46, *p*<0.05), instead of teacher-reported childhood symptoms (*d*=0.16, *p*=0.07).

For the association between neurodevelopmental conditions and ADHD persistence, although the moderating effect of the reporter (QM=6.39, *p*<0.05) and the moderating effect of the tracking period (into adolescence or adulthood) (QM=10.60, *p*<0.05) were significant. However, we found that neurodevelopmental conditions were not associated with ADHD persistence in studies using either parent-reported (*d*=0.26, *p*=0.29) or teacher-reported (*d*=0.35, *p*=0.11) childhood ADHD and comorbid symptoms. Similarly, neurodevelopmental conditions did not significantly correlate with ADHD persistence in studies tracking participants into adolescence (*d*=0.28, *p*=0.06) or into adulthood (*d*=-0.15, *p*=0.16).

The method of assessing childhood comorbidities, sex, participant age at the start of study, and reporter for adult ADHD did not significantly influence estimated associations between the persistence of ADHD and the presence of comorbidities.

### 3.4 Sensitivity analysis

As we adopted the most conservative approach to estimate unreported p values in adjusted results, a sensitivity analysis was conducted to explore whether the reduction in effect sizes in adjusted results was attributable to studies lacking specific data. Consequently, we excluded the seven studies without specific p values (Biederman et al., 1996; Tandon et al., 2011; Murray et al., 2022; Mick et al., 2011; Miranda et al., 2015; Elkins et al., 2020; Biederman et al., 2011), and reran the meta-analysis (see **Figure S4**). The adjusted results continued to indicate smaller Cohen’s d than that of unadjusted results for the association between externalizing conditions and ADHD persistence, *d*=0.12 (0.05, 0.19), consistent with the findings that included all studies. There were not enough studies to conduct the meta-analysis for internalizing conditions and neurodevelopmental conditions.

There were also four studies that used different criteria to define “persistent ADHD” (Biederman et al., 1996; Biederman et al., 2012; McAuley et al., 2017; Biederman et al., 2011). We excluded these four studies and re-ran the meta-analysis (see **Figure S5a, Figure S5b, Figure S6a, Figure S6b**). Smaller Cohen’s d values were observed in the models that included adjusted internalizing (*d*=0.09, CI=-0.05, 0.24) and externalizing (*d*=0.24, CI=0.10, 0.38) problems, compared to the models that included unadjusted internalizing (*d*=0.19, CI=0.07, 0.32) and externalizing (*d*=0.32, CI=0.20, 0.45) problems. These results were consistent with our main analysis, indicating that removing these studies did not affect the interpretation of the findings.

### 3.5 Quality of studies and publication bias

The results of our quality assessment are shown in Table 1. All studies were rated as “good” (30.8%) or “fair” (69.2%). However, the results still indicated potential publication bias in Egger’s test in the models in the internalizing (*t*=4.28, *p*<.05), externalizing (*t*=5.32, *p*<.05) and neurodevelopmental disorder (*t*=2.49, *p*<.05) models. We were unable to use the trim-and-fill method typically used to correct for potential small study bias in models (Duval & Tweedie, 2000); as it is not suitable for multilevel models.

Funnel plots (see Figures S7, S8, S9, and S10) indicated positive publication bias, with more points falling to the right of the reference line. Upon further examination of these studies, we found that the publication bias was primarily driven by studies that included teacher-reported or self-reported information.

Among the studies that used only parent-reported information (see Figure S11a, Figure S11b, Figure S12), we still found a significant unadjusted association between internalizing conditions, externalizing conditions, and ADHD persistence, with pooled Cohen’s d values of 0.18 (0.03, 0.32) and 0.48 (0.26, 0.70), respectively. After controlling for confounding variables, the association between externalizing conditions and ADHD persistence was no longer significant, with a pooled Cohen’s d value of 0.23 (−0.20, 0.67) in the only three studies remained. However, since only three studies remained after adjusting for confounders (Tandon et al., 2011; Murray et al., 2022; Tandon et al., 2011), we were unable to test for publication bias in the models of internalizing conditions or neurodevelopmental conditions and ADHD persistence that include only parent-reported information.

## 4. Discussion

We conducted a series of comprehensive meta-analyses to examine the association between childhood internalizing, externalizing, neurodevelopmental conditions and the persistence of ADHD. The unadjusted results indicated that childhood comorbid internalizing and externalizing conditions were significantly correlated with ADHD persistence. We did not find significant correlations between neurodevelopmental conditions and ADHD persistence. In the meta-analysis of studies including adjusted results, the association between externalizing conditions and ADHD persistence diminished, and the association between internalizing conditions and ADHD persistence was no longer significant. Therefore, accounting for possible confounders (e.g., sex, age, other comorbidities) can reduce the association between childhood comorbidities and ADHD persistence.

### 4.1 The association between comorbid internalizing, externalizing or neurodevelopmental conditions and ADHD persistence

The association between internalizing conditions and ADHD persistence diminished when only including studies that reported adjusted results. Children with more internalizing conditions were more likely to persist with their ADHD symptoms into later developmental stages (e.g. Hurtig et al., 2007; Mick et al., 2011; Musser et al., 2016). However, the effect size of the association between internalizing conditions and ADHD persistence is relatively low. As a result, although adding the moderating variable (e.g., sex, age, other comorbidities, etc.) did not significantly change estimated associations, it did widen confidence intervals, meaning associations became non-significant. Therefore, the role of internalizing conditions on ADHD persistence might be worth exploring again after more relevant studies are published, particularly those that report adjusted results.

Externalizing conditions were significantly associated with ADHD persistence both before and after controlling for covariates, that children with more externalizing conditions are more likely to continue experiencing ADHD symptoms. This may be due to symptomatic overlaps or shared physiological mechanisms between externalizing conditions and ADHD. The symptoms of externalizing conditions have high overlap with ADHD symptoms, such as impulsive behavior, having difficulties following rules and emotional dysregulation (Martel, Levinson, Lee, & Smith., 2017; Janssens et al., 2015; Shader, & Beauchaine, 2020). Children with ADHD and comorbid externalizing conditions typically exhibit more severe ADHD symptoms (Armstrong et al., 2015; Lapalme et al., 2018), also making it more likely for them to meet ADHD diagnostic criteria later on. Therefore, children who typically have more externalizing conditions are also more likely to meet additional diagnostic criteria for ADHD, making it more difficult for them to achieve clinical remission or symptom reduction at later developmental stages. Beyond the overlap in symptoms, previous studies have also identified shared physiological mechanisms and genetic factors between ADHD and externalizing conditions. For example, research on neural mechanisms suggests that ADHD, along with externalizing behavior problems such as CD and ODD, is associated with sensitivity to reward and a ‘disinhibitory’ deficit (Sergeant et al., 2003). Neurobiological studies have shown functional abnormalities in regions such as the prefrontal cortex, amygdala, and striatum across these conditions (e.g., Ghosh et al., 2017; Halperin & Schulz, 2006; Noordermeer et al., 2016; Van Dessel et al., 2020). Additionally, meta-analyses in genetics indicate that the genetic correlation between ADHD and externalizing conditions is 0.49 (Andersson et al., 2020). This overlap in symptoms, neural mechanisms, and genetic characteristics may contribute to the persistence of externalizing behavior problems in childhood ADHD. Besides identifying the association between externalizing conditions and ADHD persistence, another notable finding is that we observed a reduction in the effect size, albeit non-significant, of externalizing conditions on ADHD persistence after controlling for covariates. As such, the association between externalizing conditions and ADHD persistence may not be a strong direct effect, but rather the relationship may be (at least partially) explained by confounding factors.

Neurodevelopmental conditions were not significantly associated with ADHD persistence, which aligns with previous studies. In fact, most research examining the correlation between neurodevelopmental conditions and ADHD persistence has not found significant associations (Biederman et al., 1996; Chang et al., 2011; Rinsky, 2012; Willcutt et al., 2007; Chang et al., 2011). This may be due to the changing symptoms and changing genetic impact of ADHD as it develops. Although ADHD is often classified as a neurodevelopmental disorder and has high comorbidity and high genetic overlap with other neurodevelopmental conditions such as learning disabilities and autism (Crisci et al., 2021; Antshel & Russo, 2019; Andersson et al., 2020), previous research suggests that the risk genes influencing early-onset ADHD (e.g., genes related to the dopamine system) may differ from those influencing later-stage ADHD or persistent ADHD (Palladino et al., 2019). This could result in neurodevelopmental conditions having high comorbidity and genetic overlap with early-onset ADHD (Ronald et al., 2008; Daucourt et al., 2020), but these disorders may not necessarily be associated with the risk of ADHD later in development.

Additionally, regarding the impact of specific types of comorbidities on ADHD persistence, our findings are consistent with our hypothesis and the overall analysis of externalizing disorders’ effects on ADHD persistence. Childhood ODD and CD are both associated with ADHD persistence. Similarly, in line with the overall analysis of neurodevelopmental disorders’ impact on ADHD persistence, childhood learning disabilities are not related to ADHD persistence. However, when it comes to specific internalizing problems, results indicate that childhood anxiety, as an internalizing condition, significantly predicts ADHD persistence, whereas childhood depression does not. This finding suggest a differential role of anxiety and depression in predicting ADHD persistence. However, the difference in effect sizes between anxiety and depression in predicting ADHD persistence is actually quite small, and confidence intervals suggest that the heterogeneity of studies examining the relationship between childhood depression and ADHD persistence is greater than that of studies investigating childhood anxiety and ADHD persistence. Therefore, the presence or absence of statistical significance does not necessarily imply entirely distinct underlying mechanisms for the two conditions. It may simply reflect that the heterogeneity of depressive symptoms is greater than that of anxiety symptoms.

Overall, children with comorbid internalizing and externalizing conditions are more likely to have persistent ADHD symptoms, although adjusting for confounders may weaken the association between them. These findings have important practical implications. For instance, regular follow-ups may be especially relevant for children with ADHD and comorbid conditions. Additionally, this has implications for education professionals, who may need to recognize that children with ADHD and comorbid conditions might require more long-term support, as their symptoms are more likely to persist.

### 4.2 The role of moderators

The reporter of childhood symptoms was a significant moderating variable in the association between ADHD persistence and externalizing conditions, with a significant association found only for externalizing behavior problems reported by parents. This result supports the idea that parent-reported and teacher-reported symptoms might reflect different aspects of a child’s behavior, similar to previous findings (Santos et al., 2020; Youngstrom, 2000). Many externalizing conditions are observed less frequently in school settings, which may lead to teachers’ reports not fully capturing the child’s behavior (King et al., 2018). Additionally, the issue of common informant bias should be considered. In many studies where childhood symptoms were reported by teachers, ADHD symptoms in later stages were assessed through parent or self-reports (e.g., Agnew-Blais et al., 2016; Cheung et al., 2015). In contrast, studies where childhood symptoms were reported by parents often continued to rely on parent reports in later stages (e.g., Murray et al., 2022; Biederman et al., 2011; Lecendreux et al., 2015). Therefore, the observed influence of parent-reported childhood externalizing symptoms on ADHD persistence may partly stem from common informant bias. In future research, findings should be interpreted in the context of who reported the symptoms, as it is likely to influence the results. Furthermore, as more studies in this field using different informants in childhood and adulthood are published, future research should further explore whether the conclusions of this study can be replicated across different informants.

### 4.3 Publication bias

The Egger’s test indicated the possibility of publication bias in studies where childhood symptoms were reported by teachers, suggesting that studies with significant results were more likely to be published. This could lead to an inflated overall effect size. After removing studies based on teacher reports, the predictive effect of childhood externalizing behaviors on ADHD persistence remained in the unstandardized results but disappeared in the standardized results. However, since there are few adjusted studies, they may not fully represent the lack of impact of adjusted childhood comorbid externalizing problems on ADHD persistence. Therefore, whether adjusted externalizing problems still have a significant effect on ADHD persistence warrants further investigation. Due to the limited number of studies, we were unable to test the impact of publication bias on other comorbid conditions. As more studies in this field are published in the future, further investigations will be needed to examine the influence of reporter-related publication bias on research outcomes.

### 4.4 Strengths and weaknesses

Building on Caye’s study, this research further examines the association between three types of ADHD comorbidities—internalizing, externalizing, and neurodevelopmental disorders—and ADHD persistence, while also assessing potential covariates in this association. Specifically, compared to Caye’s study, the number of included studies for each comorbidity type is much larger in the current study. As a result, certain associations that were not significant in Caye’s research, such as the predictive effect of ODD on ADHD persistence, reached statistical significance in our analysis. On the other hand, we found that MDD, which was a significant predictor of ADHD persistence in Caye’s study, was no longer significant in our findings. This may be due to the inclusion of a larger number of studies in our analysis, which increased the heterogeneity of results and widened the confidence interval, leading to a loss of significance. Additionally, our definition of depression is broader than that used in Caye’s study, suggesting that different types of emotional problems may vary in their predictive power for ADHD persistence. Overall, building on previous research, The current study further deepen our understanding of the influence of childhood ADHD comorbidities on ADHD persistence.

Our results still should be interpreted in light of some limitations. Firstly, we did not specify a particular time range for ADHD persistence from childhood; we only required ADHD measurements at two time points. In the future, when more studies become available, meta-analyses could be conducted to examine ADHD persistence from childhood to adolescence and from childhood to adulthood, as well as exploring which comorbidities may affect these transitions. Secondly, we still faced the issue of a limited number of studies when conducting meta-analyses on categorized and specific conditions. Particularly after adding covariates, some groups had insufficient sample sizes, which could potentially affect the assessment of the moderation effects of covariates. Thirdly, the assessment of the covariate “childhood reporter” might be influenced by shared informant bias, since the ADHD symptoms at the second time point were most often reported by parents or the participants, and rarely by teachers. Childhood comorbid symptoms reported by parents are more likely to be associated with the persistence of parent-reported ADHD compared to those reported by teachers. Lastly, we observed that including studies with other sources of reporters might introduce publication bias, publication bias is more likely to arise for studies that include other sources of reporters (eg teachers rather than parent or self-report). This requires further investigation as more studies become available.

## 5. Conclusion

This study focused on the association between childhood comorbidities and ADHD persistence, and it was found that internalizing and externalizing conditions were significantly associated with the persistence of childhood ADHD in analyses unadjusted for confounding. However, this association decreased or disappeared after controlling for covariates. The association between ADHD persistence and internalizing or externalizing conditions may be entirely or partly due to confounding by covariates. Comorbid neurodevelopmental conditions were not associated with ADHD persistence. Further examination of the moderating effects of possible covariates revealed that the reporter of childhood symptoms moderated the association between childhood externalizing conditions and ADHD persistence. Specifically, the association between ADHD persistence and externalizing conditions was found only in studies with parent-reported data. This study explored the relationship between various comorbidities and ADHD persistence, which has important clinical implications for the intervention and management of ADHD and its comorbid conditions.

## Data Availability

All data used in the article is secondary data taken from published articles. Data may be requested from the corresponding author

## Acknowledgements

YY is supported by a China Scholarship Council; F.M. is supported by the UKRI Economic and Social Research Council (ref number: 2613456). T.S. is supported by a Wellcome Trust Sir Henry Wellcome fellowship (grant 218641/Z/19/Z). The positions of T.A.M. and Y.I.A. were funded by a Wellcome Trust Senior Research Fellowship awarded to T.A.M. (grant number 220382/Z/20/Z).

## Key points

- Children diagnosed with ADHD and other comorbid mental health conditions show a greater tendency for their ADHD symptoms to persist into later developmental stages.
- We conducted a systematic review and meta-analysis including 26 studies to investigate the extent to which specific childhood comorbidities predict the persistence of childhood ADHD into later developmental stages.
- Childhood comorbid externalizing and, to a lesser extent, internalizing conditions were associated with the persistence of ADHD, but this association may be partially due to confounders.
- Our results have implications for education professionals, who may need to recognize that children with ADHD and comorbid conditions might require more long-term support.

