## Supporting information for "The role of comorbid childhood mental health conditions in the persistence of ADHD symptoms: Systematic review and Meta-analysis"

**Appendix S1**. Search strategies
**Figure S1a**. The predictive effect of depression on ADHD persistence: Unadjusted
**Figure S1b**. The predictive effect of anxiety on ADHD persistence: Unadjusted
**Figure S1c**. The predictive effect of depression on ADHD persistence: Adjusted
**Figure S1d**. The predictive effect of anxiety on ADHD persistence: Adjusted
**Figure S2a.** The predictive effect of oppositional defiant disorder on ADHD persistence: Unadjusted
**Figure S2b.** The predictive effect of conduct disorder on ADHD persistence: Unadjusted
**Figure S2c.** The predictive effect of oppositional defiant disorder on ADHD persistence: Adjusted
**Figure S2d.** The predictive effect of conduct disorder on ADHD persistence: Adjusted
**Figure S2e.** The predictive effect of aggression on ADHD persistence: Adjusted
**Figure S3.** The predictive effect of learning disorder on ADHD persistence: Unadjusted
**Figure S4**. The predictive effect of externalizing on ADHD persistence: Adjusted (after excluding the studies without specific p value)
**Figure S5a**. The predictive effect of externalizing on ADHD persistence: Unadjusted (after excluding the studies using different criterion of ADHD persistence)
**Figure S5b**: The predictive effect of externalizing on ADHD persistence: Adjusted (after excluding the studies using different criterion of ADHD persistence)
**Figure S6a**: The predictive effect of internalizing on ADHD persistence: Unadjusted results (after excluding the studies using different criterion of ADHD persistence)
**Figure S6b**. The predictive effect of internalizing on ADHD persistence: Adjusted results (after excluding the studies using different criterion of ADHD persistence)
**Figure S7.** The funnel plot of the internalizing conditions on ADHD persistence
**Figure S8.** The funnel plot of the externalizing conditions on ADHD persistence
**Figure S9**. The funnel plot of the neurodevelopmental conditions on ADHD persistence
**Figure S10.** The funnel plot of the externalizing conditions on ADHD persistence (use only parent-reported information)
**Figure S11a**. The predictive effect of externalizing on ADHD persistence: Unadjusted (use only parent-reported information)
**Figure S11b**. The predictive effect of externalizing on ADHD persistence: Adjusted (use only parent-reported information)
**Figure S12**. The predictive effect of internalizing on ADHD persistence: Unadjusted (use only parent-reported information)

**Appendix S1**. Search strategies

**Population**: child* OR boys OR girls OR kindergarten OR students OR primary OR youth OR young OR teenager OR adolescen* OR youngster

**Exposure**: comorbid* OR emotional problems OR internalizing OR depressi* OR anxi* OR neurodevelopment OR neuroatypical OR ODD OR oppositional defiant disorder OR autism OR learning disorder OR dyslexia OR externalizing OR psychopatholog* OR mental disorders OR wellbeing OR psychological disorders OR adjustment disorders OR mental health

**Type of study**: Predict* OR associate* OR detect* OR forecast* OR longitudinal OR follow-up OR prospective

**Outcome**: (persistent adj3 ADHD) OR (ADHD adj3 persistence) OR (ADHD adj3 remission) OR (continu* ADHD symptoms) OR (ADHD adj3 trajectory*) OR (remitted adj3 ADHD) OR (unremitted adj3 ADHD) OR (grow out of ADHD) OR (ADHD adj3 outcomes) OR (persistent adj3 inattent*) OR (inattent* adj3 persistence) OR (inattent* adj3 remission) OR (continu* inattent* symptoms) OR (inattent* adj3 trajector*) OR (remitted adj3 inattent*) OR (unremitted adj3 inattent*) OR (grow out of inattent*) OR (inattent* adj3 outcomes) OR (persistent adj3 hyperactiv*) OR (hyperactiv* adj3 persistence) OR (hyperactiv* adj3 remission) OR (continu* hyperactiv* symptoms) OR (hyperactiv* adj3 trajector*) OR (remitted adj3 hyperactiv*) OR (unremitted adj3 hyperactiv*) OR (grow out of hyperactiv*) OR (hyperactiv* adj3 outcomes) OR (ADHD adj3 stability) OR (inattent* adj3 stability) OR (hyperactiv* adj3 stability)

**Figure S1a**. The predictive effect of depression on ADHD persistence: Unadjusted

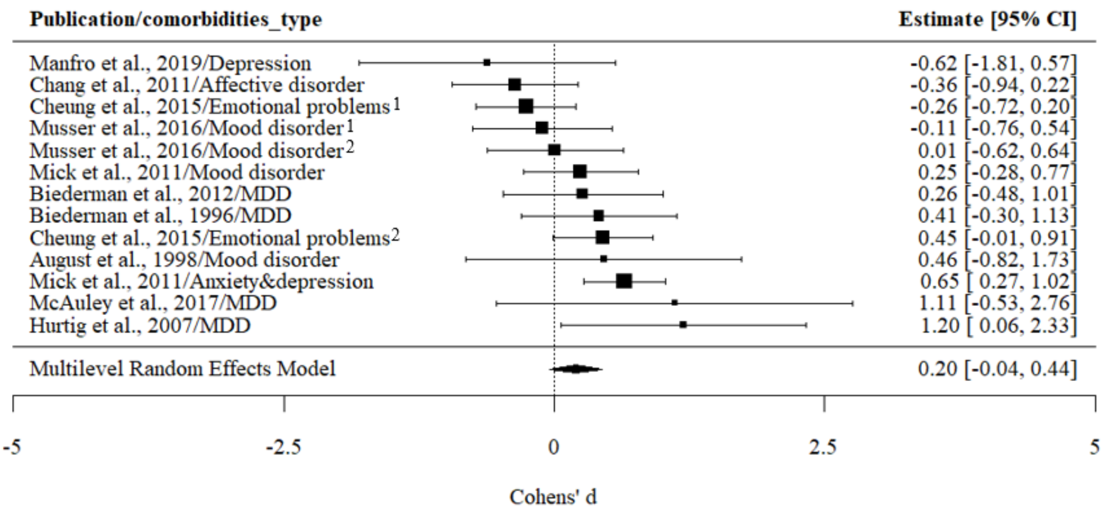


**Note**: Cheung et al., 2015/Emotional problems[1]: emotional problems were reported by teachers; Cheung et al., 2015/Emotional problems[2]: emotional problems were reported by parents; Musser et al., 2016/Mood disorder[1]: mood disorder was reported by teachers; Musser et al., 2016/Mood disorder[2]: mood disorder was reported by parents.

**Figure S1b**. The predictive effect of anxiety on ADHD persistence: Unadjusted

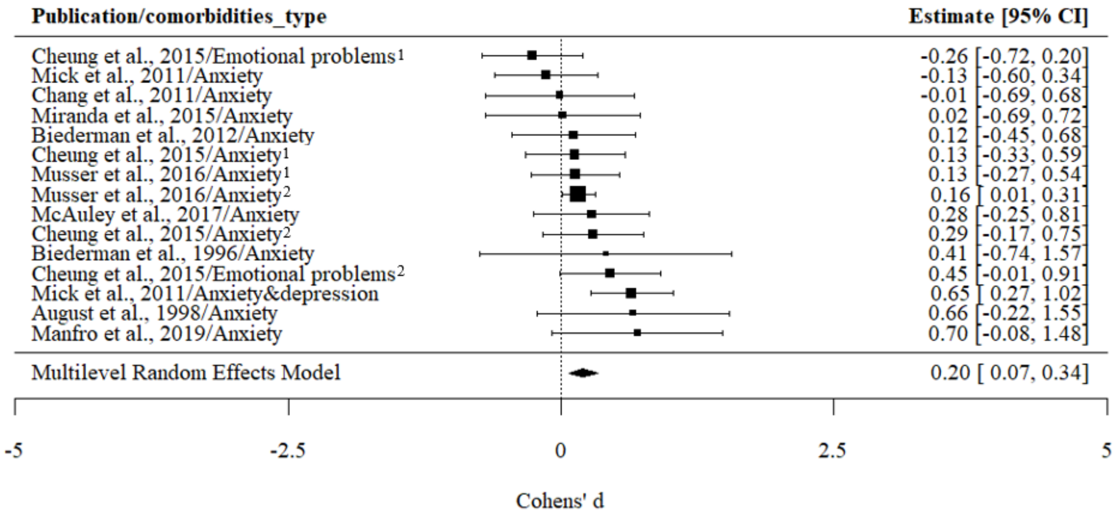


**Note**: Cheung et al., 2015/Emotional problems[1]: emotional problems were reported by teachers; Cheung et al., 2015/Emotional problems[2]: emotional problems were reported by parents. Cheung et al., 2015/Anxiety[1]: anxiety was reported by teachers; Cheung et al., 2015/Anxiety[2]: anxiety was reported by parents; Musser et al., 2016/Anxiety[1]: anxiety was reported by teachers; Musser et al., 2016/Anxiety[2]: anxiety was reported by parents.

**Figure S1c**. The predictive effect of depression on ADHD persistence: Adjusted

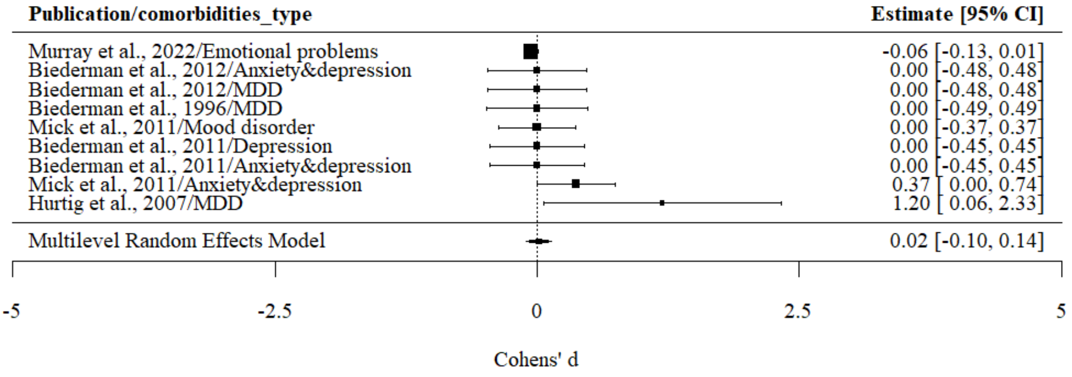


**Figure S1d**. The predictive effect of anxiety on ADHD persistence: Adjusted

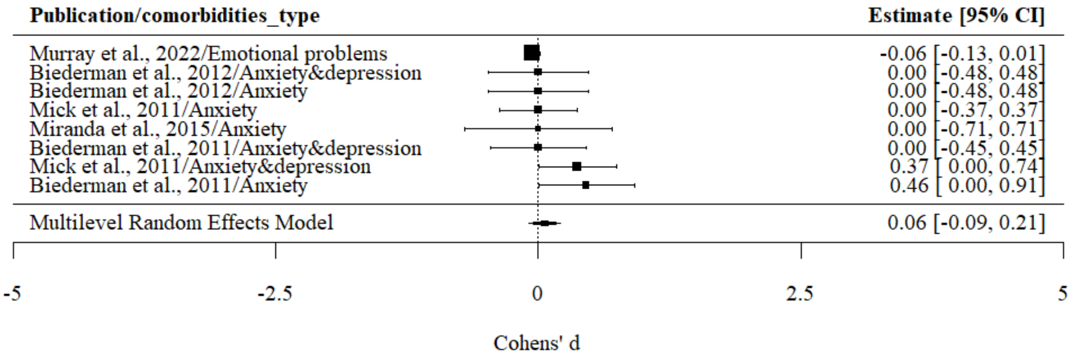

**Figure S2a.** The predictive effect of oppositional defiant disorder on ADHD persistence: Unadjusted

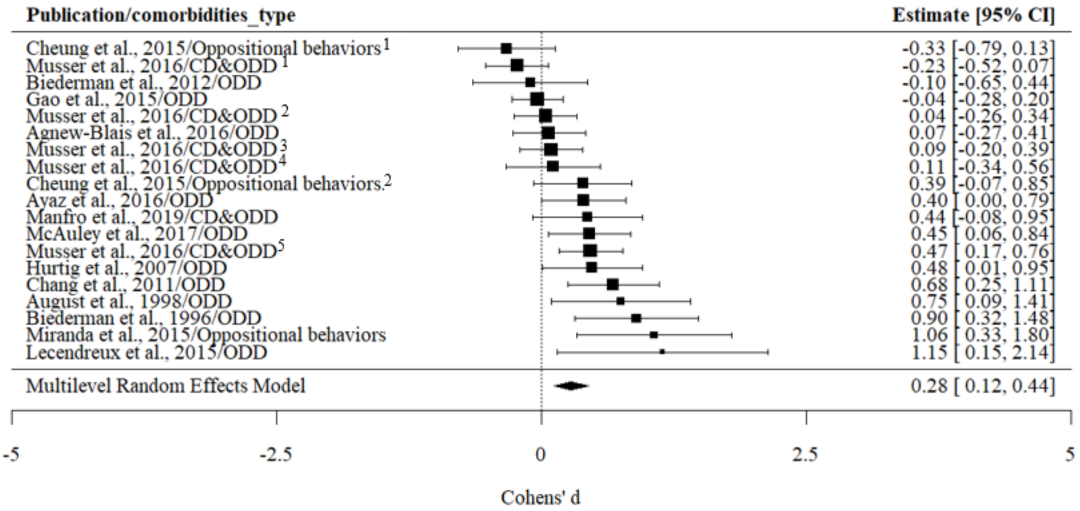


**Note**: ODD, oppositional defiant disorder; CD, conduct disorder.

Cheung et al., 2015/Oppositional behaviors[1]: oppositional behaviors were reported by teachers; Cheung et al., 2015/Oppositional behaviors[2]: oppositional behaviors were reported by parents. Musser et al., 2012/CD&ODD[1]: CD&ODD was reported by teachers, and ADHD was reported by teachers. Musser et al., 2012/CD&ODD[2]: CD&ODD were reported by teachers, and ADHD was reported by parents. Musser et al., 2012/CD&ODD[3]: CD&ODD were reported by parents, and ADHD was reported by teachers. Musser et al., 2012/CD&ODD[4]: CD&ODD were assessed by diagnostic team, and ADHD was reported by teachers. Musser et al., 2012/CD&ODD[5]: CD&ODD were reported by parents, and ADHD was reported by parents.

**Figure S2b.** The predictive effect of conduct disorder on ADHD persistence: Unadjusted

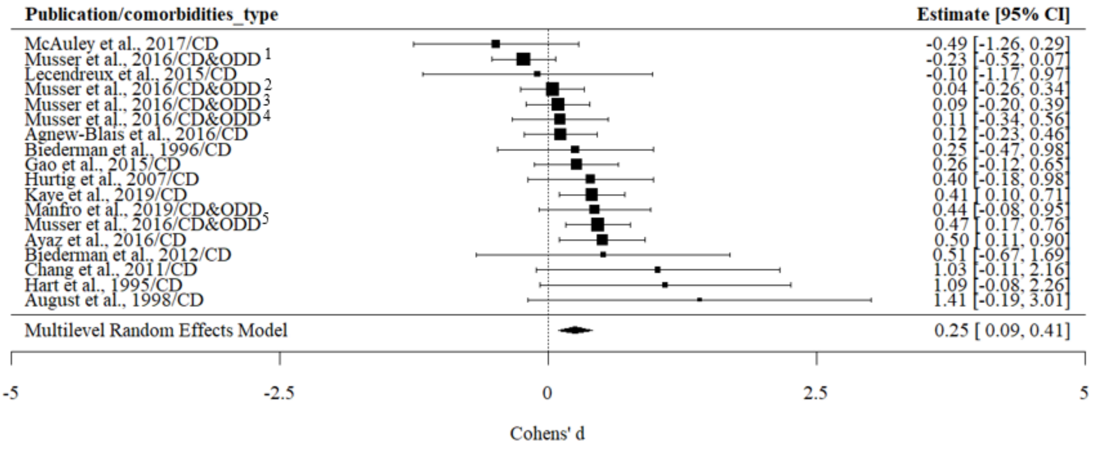


**Note**: ODD, oppositional defiant disorder; CD, conduct disorder.

Musser et al., 2012/CD&ODD[1]: CD&ODD was reported by teachers, and ADHD was reported by teachers. Musser et al., 2012/CD&ODD[2]: CD&ODD were reported by teachers, and ADHD was reported by parents. Musser et al., 2012/CD&ODD[3]: CD&ODD were reported by parents, and ADHD was reported by teachers. Musser et al., 2012/CD&ODD[4]: CD&ODD were assessed by diagnostic team, and ADHD was reported by teachers. Musser et al., 2012/CD&ODD[5]: CD&ODD were reported by parents, and ADHD was reported by parents.

**Figure S2c.** The predictive effect of oppositional defiant disorder on ADHD persistence: Adjusted

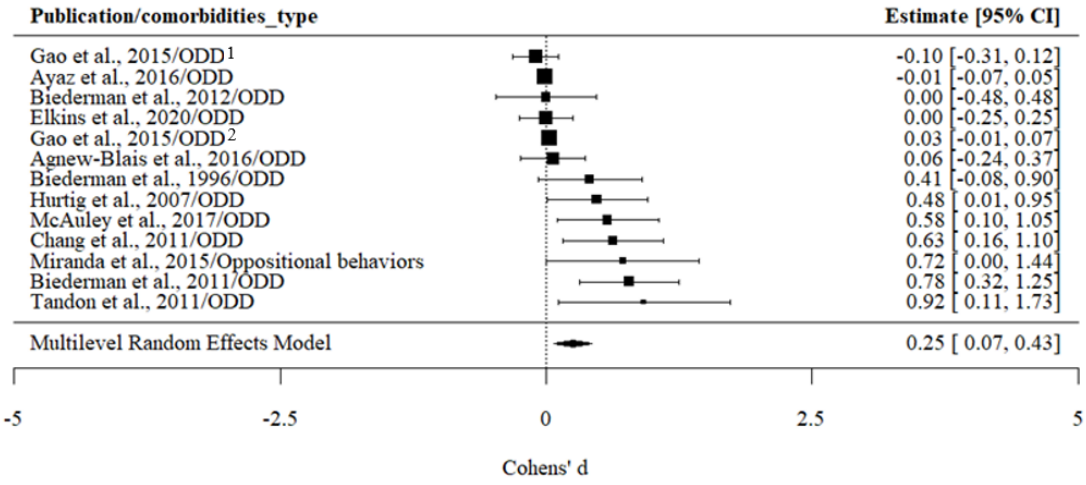


**Note:** ODD, oppositional defiant disorder

Gao et al., 2015/ODD[1]: ODD was included as a categorical indicator; Gao et al., 2015/ODD[2]: ODD was included as a quantitative traits.

**Figure S2d.** The predictive effect of conduct disorder on ADHD persistence: Adjusted

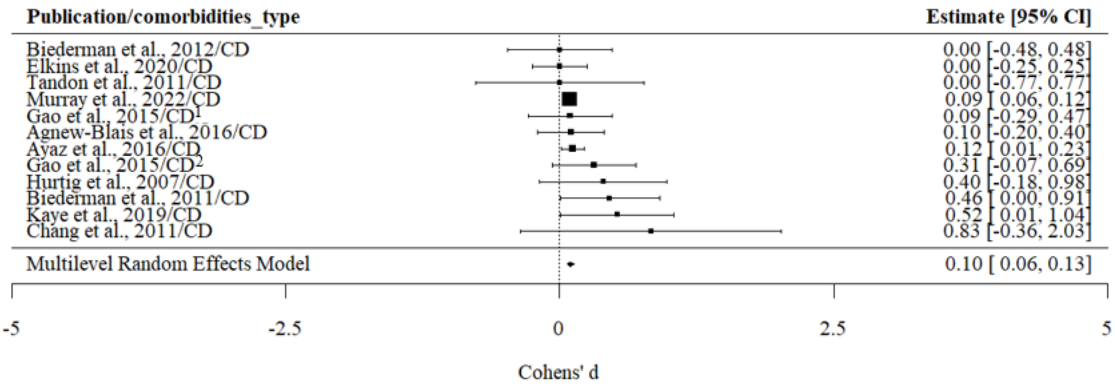


**Note:** Gao et al., 2015/CD[1]: CD was included as a categorical indicator; Gao et al., 2015/CD[2]: CD was included as a quantitative traits.

**Figure S2e.** The predictive effect of aggression on ADHD persistence: Adjusted

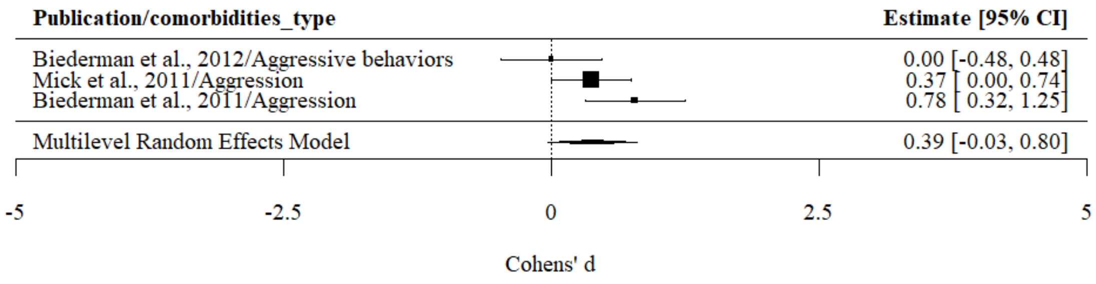


**Figure S3.** The predictive effect of learning disorder on ADHD persistence: Unadjusted

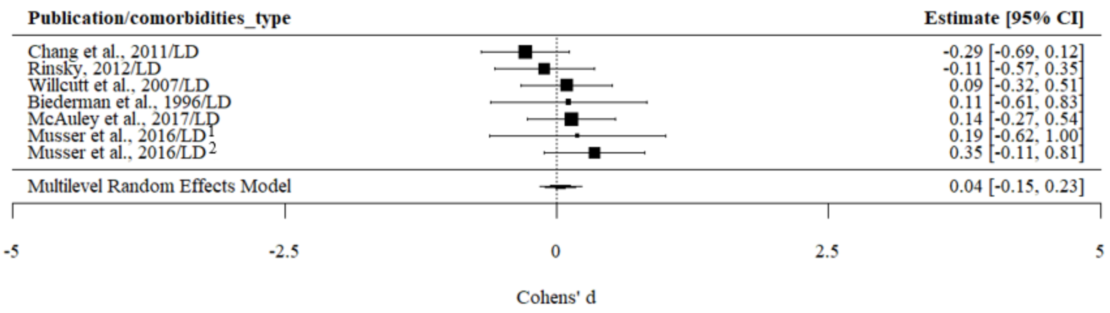


**Note:** Musser et al., 2016/LD[1]: LD was reported by teachers; Musser et al., 2016/LD[2]: LD was reported by parents.

### Sensitive analysis

**Figure S4**. The predictive effect of externalizing on ADHD persistence: Adjusted (after excluding the studies without specific p value)

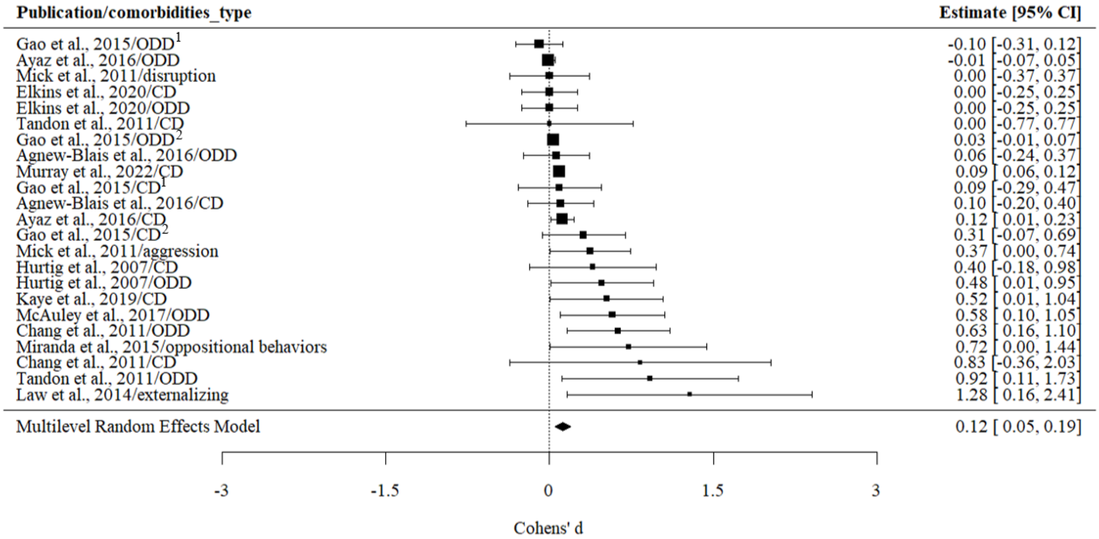


**Note**: Gao et al., 2015/ODD[1]: ODD was included as a categorical indicator; Gao et al., 2015/ODD[2]: ODD was included as a quantitative traits. Gao et al., 2015/CD[1]: CD was included as a categorical indicator; Gao et al., 2015/ODD[2]: ODD was included as a quantitative traits.

**Figure S5a**. The predictive effect of externalizing on ADHD persistence: Unadjusted (after excluding the studies using different criterion of ADHD persistence)

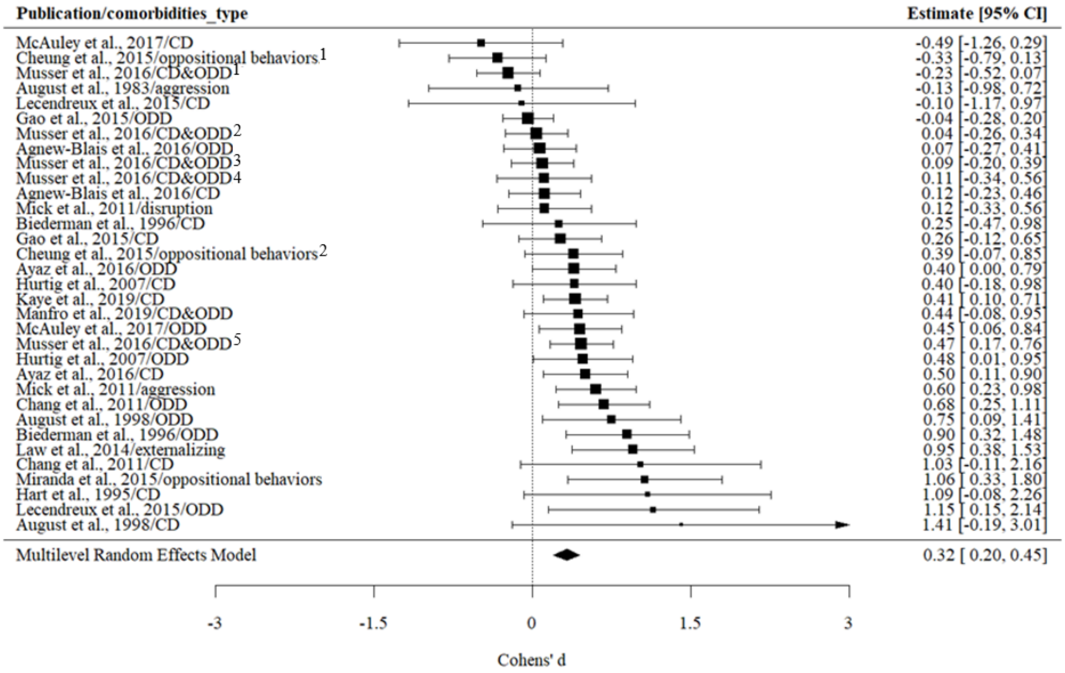


**Note**: Cheung et al., 2015/Oppositional behaviors[1]: oppositional behaviors were reported by teachers; Cheung et al., 2015/Oppositional behaviors[2]: oppositional behaviors were reported by parents. Musser et al., 2012/CD&ODD[1]: CD&ODD was reported by teachers, and ADHD was reported by teachers. Musser et al., 2012/CD&ODD[2]: CD&ODD were reported by teachers, and

ADHD was reported by parents. Musser et al., 2012/CD&ODD[3]: CD&ODD were reported by parents, and ADHD was reported by teachers. Musser et al., 2012/CD&ODD[4]: CD&ODD were assessed by diagnostic team, and ADHD was reported by teachers. Musser et al., 2012/CD&ODD[5]: CD&ODD were reported by parents, and ADHD was reported by parents.

**Figure S5b**: The predictive effect of externalizing on ADHD persistence: Adjusted (after excluding the studies using different criterion of ADHD persistence)

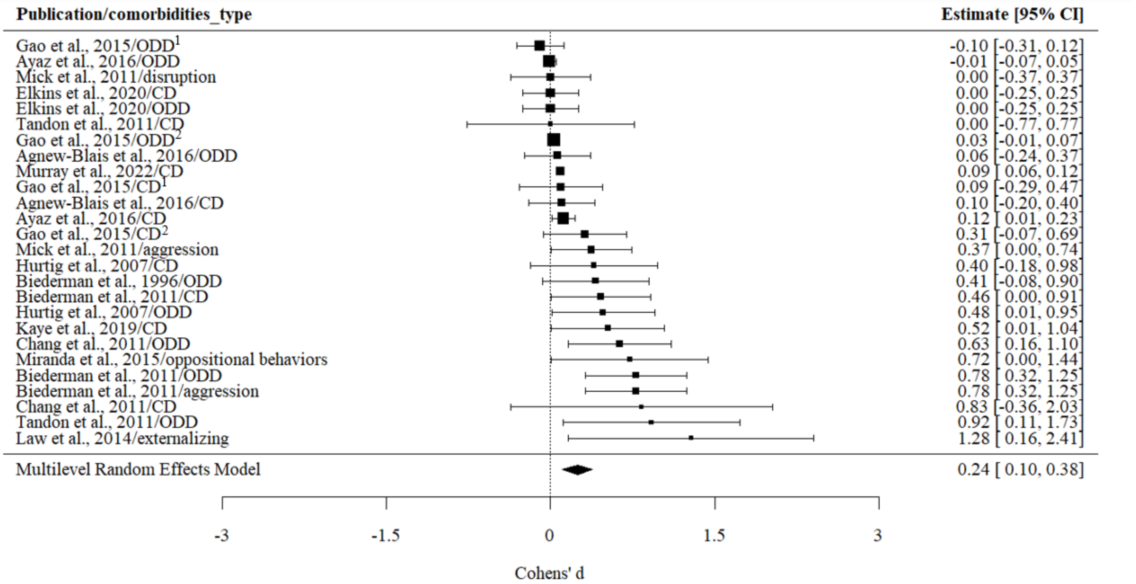


**Note**: Gao et al., 2015/ODD[1]: ODD was included as a categorical indicator; Gao et al., 2015/ODD[2]: ODD was included as a quantitative traits. Gao et al., 2015/CD[1]: CD was included as a categorical indicator; Gao et al., 2015/ODD[2]: ODD was included as a quantitative traits.

**Figure S6a**: The predictive effect of internalizing on ADHD persistence: Unadjusted results (after excluding the studies using different criterion of ADHD persistence)

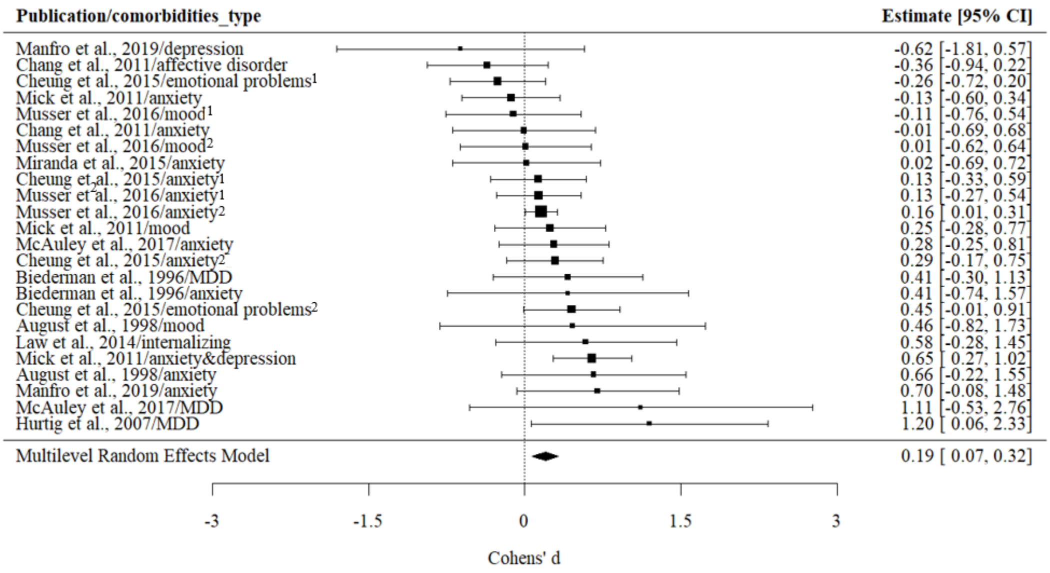


**Note**: Cheung et al., 2015/Emotional problems[1]: emotional problems were reported by teachers; Cheung et al., 2015/Emotional problems[2]: emotional problems were reported by parents. Cheung et al., 2015/Anxiety[1]: anxiety was reported by teachers; Cheung et al., 2015/Anxiety[2]: anxiety was reported by parents; Musser et al., 2016/Anxiety[1]: anxiety was reported by teachers; Musser et al., 2016/Anxiety[2]: anxiety was reported by parents. Musser et al., 2016/Mood disorder[1]: mood disorder was reported by teachers; Musser et al., 2016/Mood disorder[2]: mood disorder was reported by parents.

**Figure S6b**. The predictive effect of internalizing on ADHD persistence: Adjusted results`(after excluding the studies using different criterion of ADHD persistence)

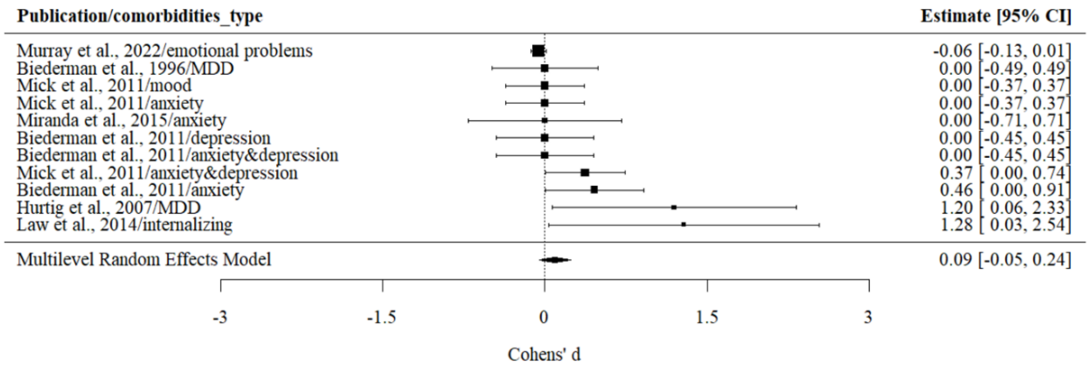

**Publication bias**

Funnel plot (using all the studies)

**Figure S7.** The funnel plot of the internalizing conditions on ADHD persistence

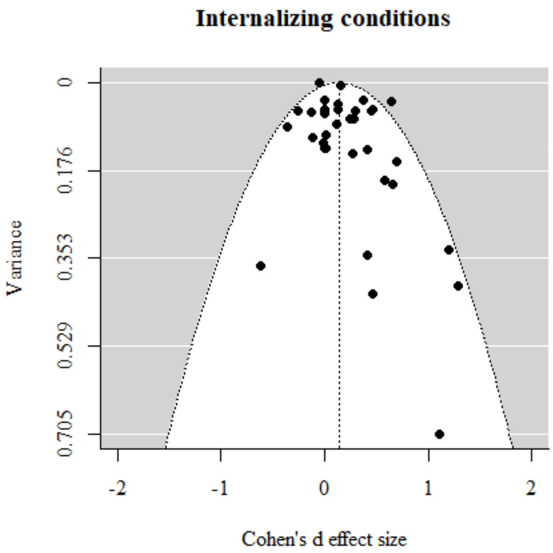


**Figure S8.** The funnel plot of the externalizing conditions on ADHD persistence

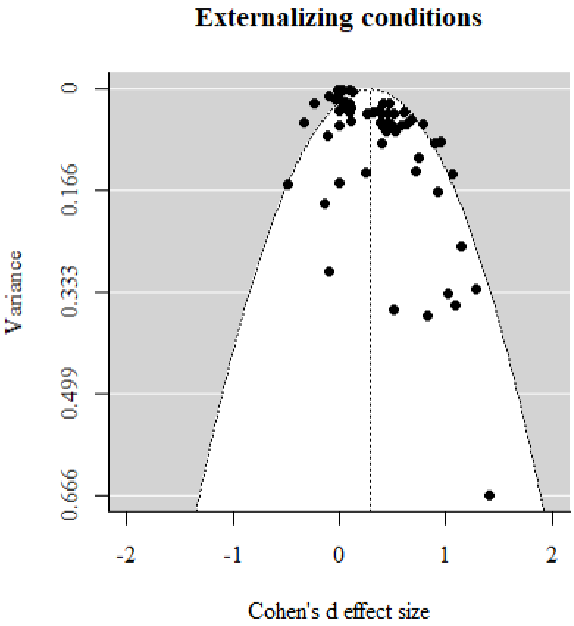

**Figure S9**. The funnel plot of the neurodevelopmental conditions on ADHD persistence

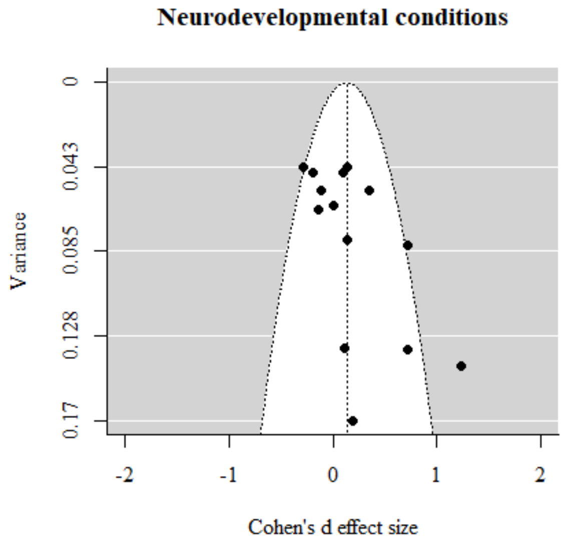


Funnel plot (use only parent-reported information)

**Figure S10.** The funnel plot of the externalizing conditions on ADHD persistence (use only parent-reported information)

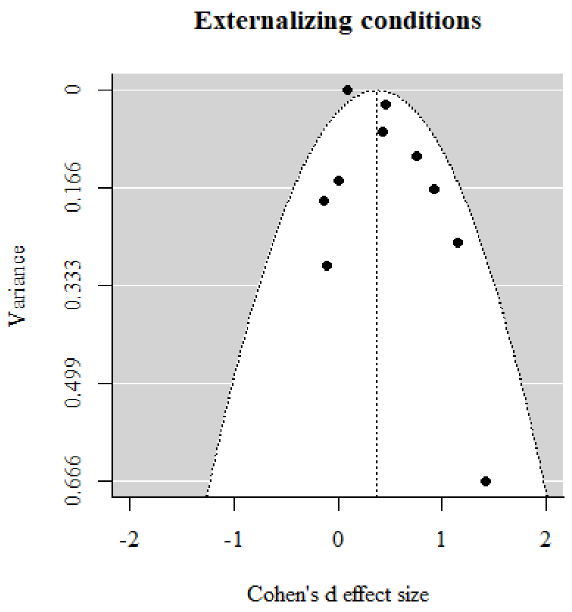

**Meta-analysis results of studies based only on parent-reported information**

**Figure S11a**. The predictive effect of externalizing on ADHD persistence: Unadjusted (use only parent-reported information)

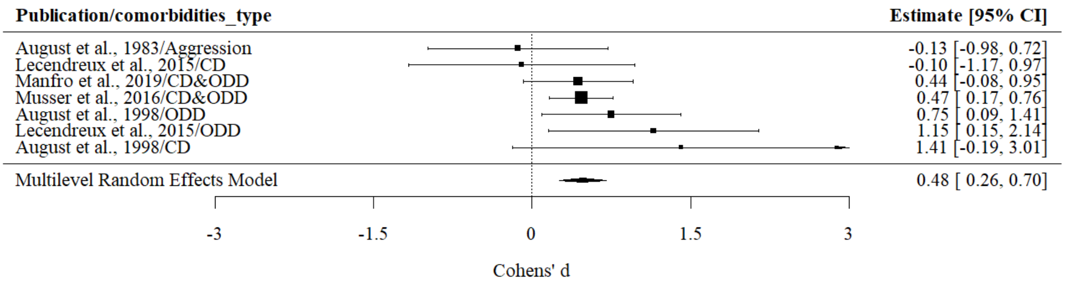


**Figure S11b**. The predictive effect of externalizing on ADHD persistence: Adjusted (use only parent-reported information)

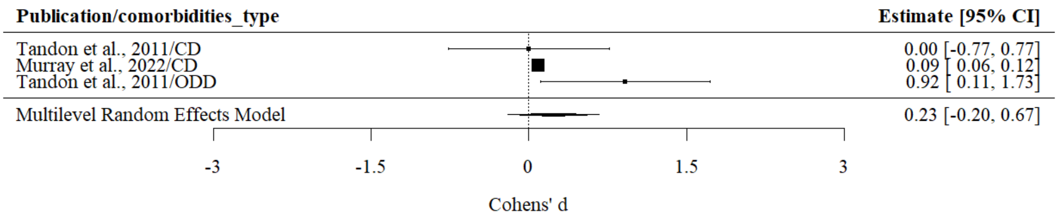


**Figure S12**. The predictive effect of internalizing on ADHD persistence: Unadjusted (use only parent-reported information)

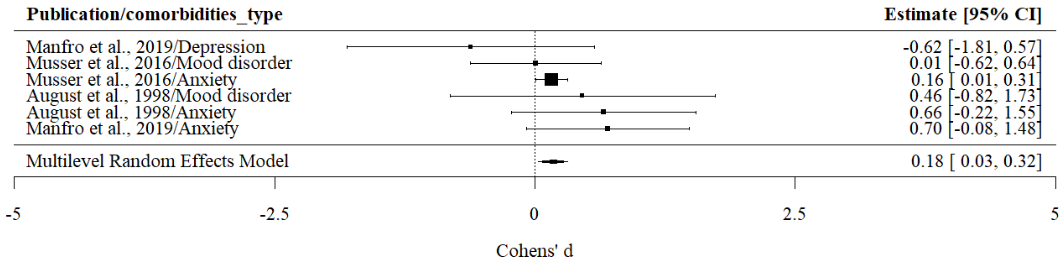
